# High-Intensity Interval Training Improves physical morphology, Cardiopulmonary Fitness and Metabolic Risk Indicators of Cardiovascular Disease in Children and Adolescents: A Systematic Review and Meta-Analysis

**DOI:** 10.1101/2022.07.11.22277515

**Authors:** Menjie, Zou Shuangling, Majia, Xiang chenmin, Li Shufeng, Wang Junli

## Abstract

**Objective:** To systematically evaluate the safety and efficacy of high-intensity interval training in children and adolescents.

**Design:** Systematic Review and Meta-Analysis

**Methods:** Eight databases were searched. Descriptive analysis of the efficacy and safety of high-intensity interval training on body shape, cardiorespiratory fitness, and metabolic risk markers for cardiovascular disease between children and adolescents. Subgroup analysis was performed with age, participants, intervention time, and exercise frequency as covariates.

**Results:** 47 studies included 2995 children and adolescents. Meta-analysis results showed that high-intensity interval training significantly improved cardiorespiratory fitness indicators (VO_2max_, SBP, DBP and HR_max_) and cardiovascular disease metabolic risk indicators (TC, HDL-C). HIIT had no significant effect on body shape indicators (BMI, BF% and WC) and some cardiometabolic indicators (TG and LDL-C).

**Conclusion:** Current evidence shows insufficient evidence that high-intensity interval training with intermittent running as the main form of exercise improves body shape indicators in children and adolescents. Nevertheless, it deserves to recommend for the purpose of improving cardiorespiratory fitness and reducing the metabolic risk of cardiovascular disease.

## INTRODUCTION

In 2018, the World Health Organization (WHO) conducted a summary analysis of 16 million children and adolescents in 146 countries and regions based on 298 population surveys and pointed out that 85% of girls and 78% of boys in the world did not meet the WHO recommended standards, which leads to the inevitable global trend towards the insufficient physical activity of adolescents [1]. There is growing evidence that physical inactivity in children and adolescents increases the prevalence of cardiovascular disease in adulthood and affects cognitive development, social interaction and even current and future health[2, 3]. Insufficient physical activity between children and adolescents is highly correlated with metabolic diseases in adulthood, especially increasing the risk of diseases such as metabolic obesity, type 2 diabetes mellitus (T_2_DM), cardiovascular disease (CVD), and cancer[4]. To achieve the goal of reducing the insufficient rate of physical activity by 15% in 2030, it is necessary to enhance the level of physical activity of children and adolescents[5]. What we all know is that continuous aerobic exercise can increase the aerobic capacity of the body, improve the sensitivity of insulin resistance, improve the level of lipometabolism and reduce the risk of diseases caused by physical inactivity. However, aerobic exercise lasts for a long time and has a single rhythm, which makes it difficult for most people to persist. But one of the main obstacles to achieving regular physical activity for current children and adolescents is the lack of time. Therefore, it is likely to replace aerobic exercise with high-intensity interval training (HIIT), because HIIT has the advantages of low time cost, less exercise volume, easy persistence, and equivalent exercise effect to aerobic exercise. HIIT refers to the training method that is repeated multiple times at a greater than or equal to anaerobic threshold or maximal lactate steady-state intensity with incomplete recovery between each set of exercises. The body is more sensitive to the stimulation of HIIT and produces more comprehensive benefits in terms of sports ability, skeletal muscle metabolism and energy consumption.

In recent years, the comparison between HIIT and continuous aerobic exercise effect value has become a hot research topic, and the research groups are mainly obese and athletes: Obese people focus on body composition, metabolism and cardiorespiratory fitness (CRF), while athletes are primarily concerned with athletic performance and physiological adaptation during exercise[6].

HIIT research on obese children and adolescents and normal children and adolescents has gradually attracted attention and achieved certain research results. A recently published meta-analysis of HIIT targeting obese children and adolescents showed that HIIT was effective in improving cardiometabolic level, cardiopulmonary adaptability, and aerobic capacity of obese children and adolescents, but the evidence for conclusions about body composition improvement is insufficient[7]. Meta-analysis of healthy children and adolescents has shown that HIIT can effectively improve the health level of children and adolescents and cardiovascular disease risk factors[8]. Meta-analysis of young athletes shows that HIIT can improve the aerobic and anaerobic exercise ability of young athletes, and the time cost is lower. Comparing HIIT with moderate-intensity continuous training (MICT) found similar effects on body composition, blood pressure in childhood obesity[9] and greater improvements in cardiorespiratory fitness in children and adolescents. HIIT can be used as an alternative training mode of MICT to maintain cardiometabolic health and it can be applied to the management of childhood obesity. Whereas, previous studies have small sample sizes, deviation of outcome index measurement tools, language bias[8], and unclear description of exercise dose[10], especially lack subgroup analyses on the influence of pre-puberty and pubertygender[7] that affect the stability of results. Given the above, this study will systematically evaluate the effectiveness and safety of HIIT for children and adolescents, expecting to provide a scientific basis for the promotion of HIIT in children and adolescents.

## METHODS

### Protocol

A systematic review and meta-analyses were conducted in accordance with the 27 checklists applying the established guidelines of the PRISMA statement 2020[11], whose aim is to serve as a basis for reporting systematic reviews of randomized trials. Besides there is no registration review protocol for this study.

### Document retrieval strategy

The computer retrieves PubMed, The Cochrane Library, Embase, Web of science, Science Direct, CNKI, WanFang and VIP databases. In addition, research published by Google Scholar was hand-searched. Randomized controlled and non-randomized controlled trials on the health efficacy and safety of HIIT between children and adolescents were collected.The retrieval time limit was from the establishment of the database to January 1, 2022. We employed the following MeSH terms: High-Intensity Interval Training, High-Intensity Interval, High-Intensity Intermittent, Adolescence, Teenagers, randomized controlled trial, RCT, etc. Taking PubMed as an example, the specific search strategy is shown in Table 1.

**Table 1.**
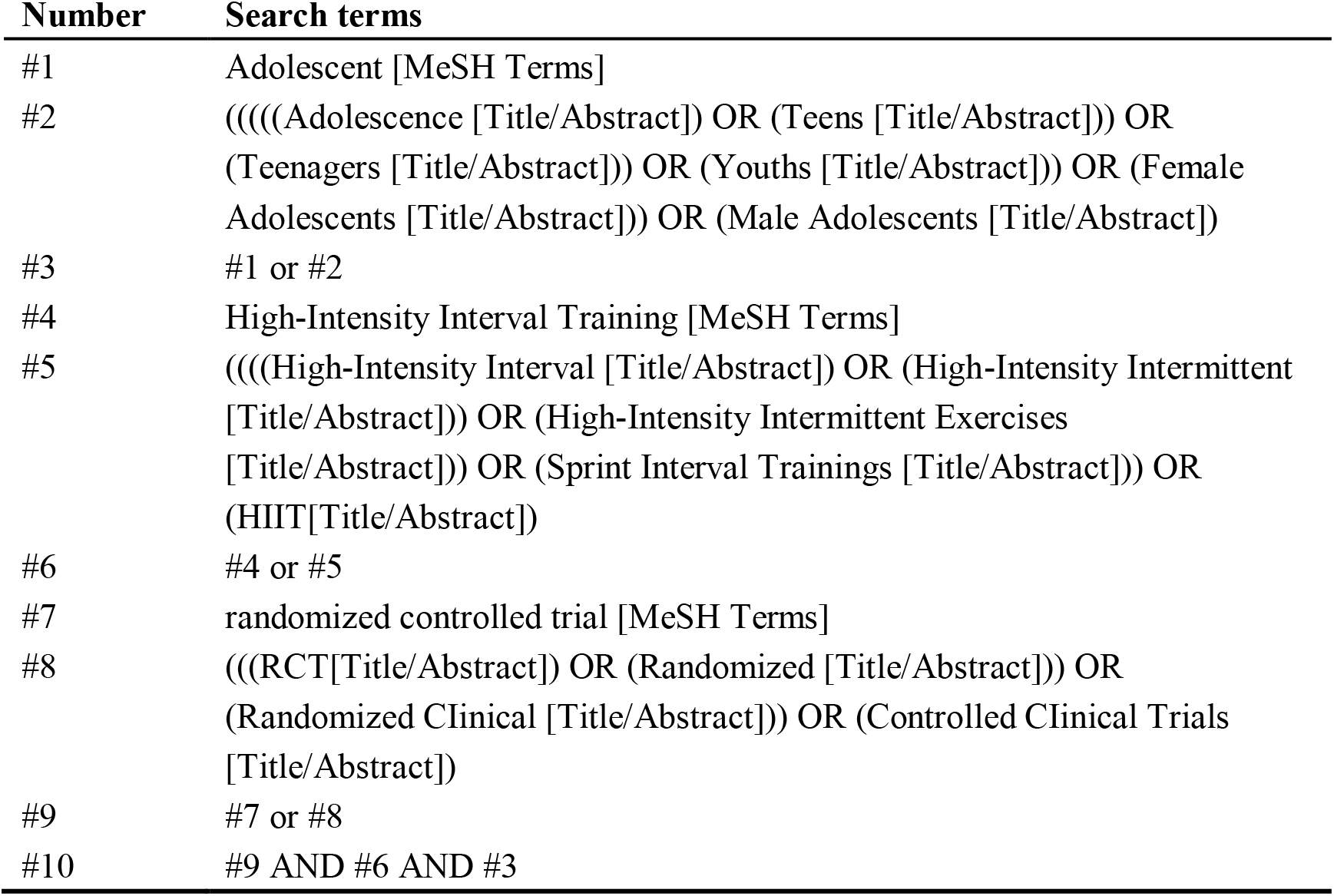
Full-search strategy for PubMed.

### Inclusion and exclusion criteria

Studies of children and adolescents were considered for the systematic review provided they met the following inclusion criteria:

- Type of study: Randomized controlled trial (RCT) or controlled trial.Participants: Children and adolescents aged 5 to 19 (Normal weight, obesity, disease, etc.).
- Interventions: The control group received no intervention. The experimental group was high-intensity interval training, and the interventions had no specific requirements except for intensity (intensity
- ≥80%HR_max_ or ≥100%aerobic speed or ≥80%VO_2max_).Outcome indexes: body shape indicators, CRF indicators and cardiovascular disease metabolic risk indicators.

Studies were excluded in the following cases:

- Not reported in Chinese or English.
- Controlled experiment before and after intervention.
- Duplicate published literature.
- Studies that could not extract important outcome data.

### Literature screening and data extraction

Two researchers (Zou Shuangling and Xiang Chenmin) independently screened the literature, extracted data and cross-checked. If there was any disagreement, it would be resolved through consultation. If additional information was required, contact the corresponding author by email. The extracted content includes:(ⅰ) the basic information of the included studies: research title, author, publication year, journal name, etc.; (ⅱ) baseline characteristics and interventions of the study subjects; (ⅲ) outcome index data and outcome index measurement methods. (ⅳ) Whether lost to follow-up, withdrawal, medical supervision measures and description of adverse reaction events, etc.

### Risk of bias and evaluation of literature quality included in the study

The bias risk assessment tools Cochrane (RoB2.0) and RevMan 5.3 independently evaluated the risk of bias in the included studies by two investigators (Menjie and Majia) and cross-checked the results.

### Statistical analysis

Meta-analysis was performed using RevMan 5.3 and Stata 15.0 software. The measurement data used mean difference (MD) as the effect analysis statistics, and each effect amount provided its 95% confidence interval (CI). Sensitivity analysis of the included studies was performed to assess data robustness, and heterogeneity magnitude was evaluated in combination with I^2^: when I^2^< 25% is low heterogeneity, I^2^=25∼50% is moderate heterogeneity, I^2^ >50% is high heterogeneity, the level of meta-analysis is set to α=0.05. If the heterogeneity between the results was not statistically significant, a fixed-effects model was used for meta-analysis; if there was statistical heterogeneity between the studies, a random-effects model was used for meta-analysis and subgroup analysis was used to analyze the sources of heterogeneity further. Publication bias is graphically assessed by Egger’s linear regression analysis.

## RESULTS

### Literature screening process and results

3,868 studies were retrieved in December 2021: 471 studies were deleted for duplication, and after title and abstract screening, 3195 studies were considered ineligible and 202 full texts were screened based on inclusion/exclusion criteria. Finally a total of 47 studies were included, and the content of the literature selection process and results are shown in Fig.1.

**Fig. 1.**
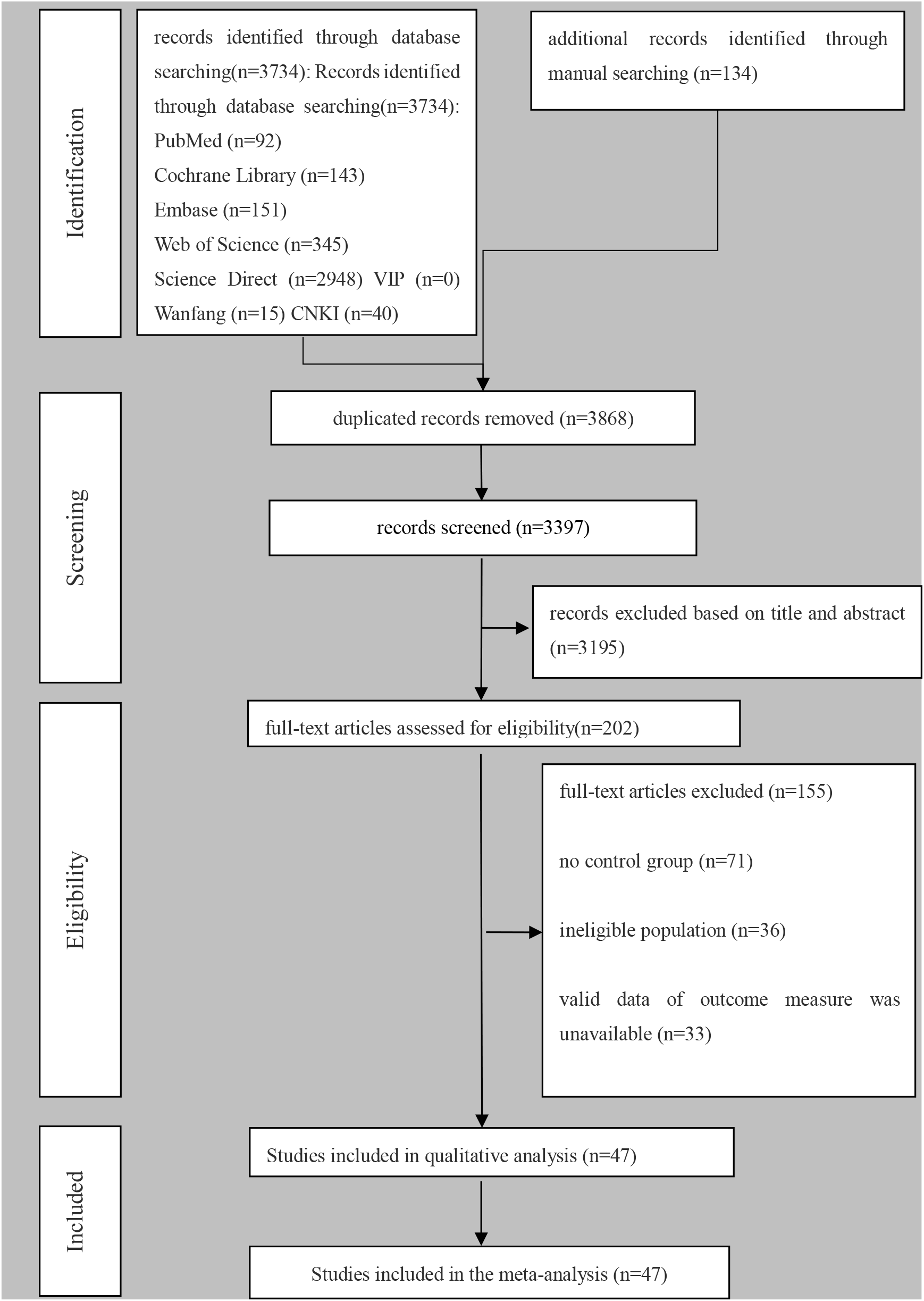
PRISMA diagram outlining the results of the screening and selection

### Incorporate basic characteristics of research

Table 2 summarizes the basic characteristics of 47 studies[12–58], which investigated a total of 2995 subjects (HIIT group: 1749, control group: 1246). Among them, there were 1165 boys (38.90%), 1156 girls (38.60%) and 696 girls (23.24%) who did not mention gender. There were 2328 children (77.73%) and 667adolescents (22.27%). 438 were overweight/obese (14.62%); 106 athletes (3.54%); 433 sick children and adolescents (14.46%); There were 26 studies with medical supervision description (55.32%), 0 studies without medical supervision and 21 studies without detailed description (44.68%).

**Table 2.**
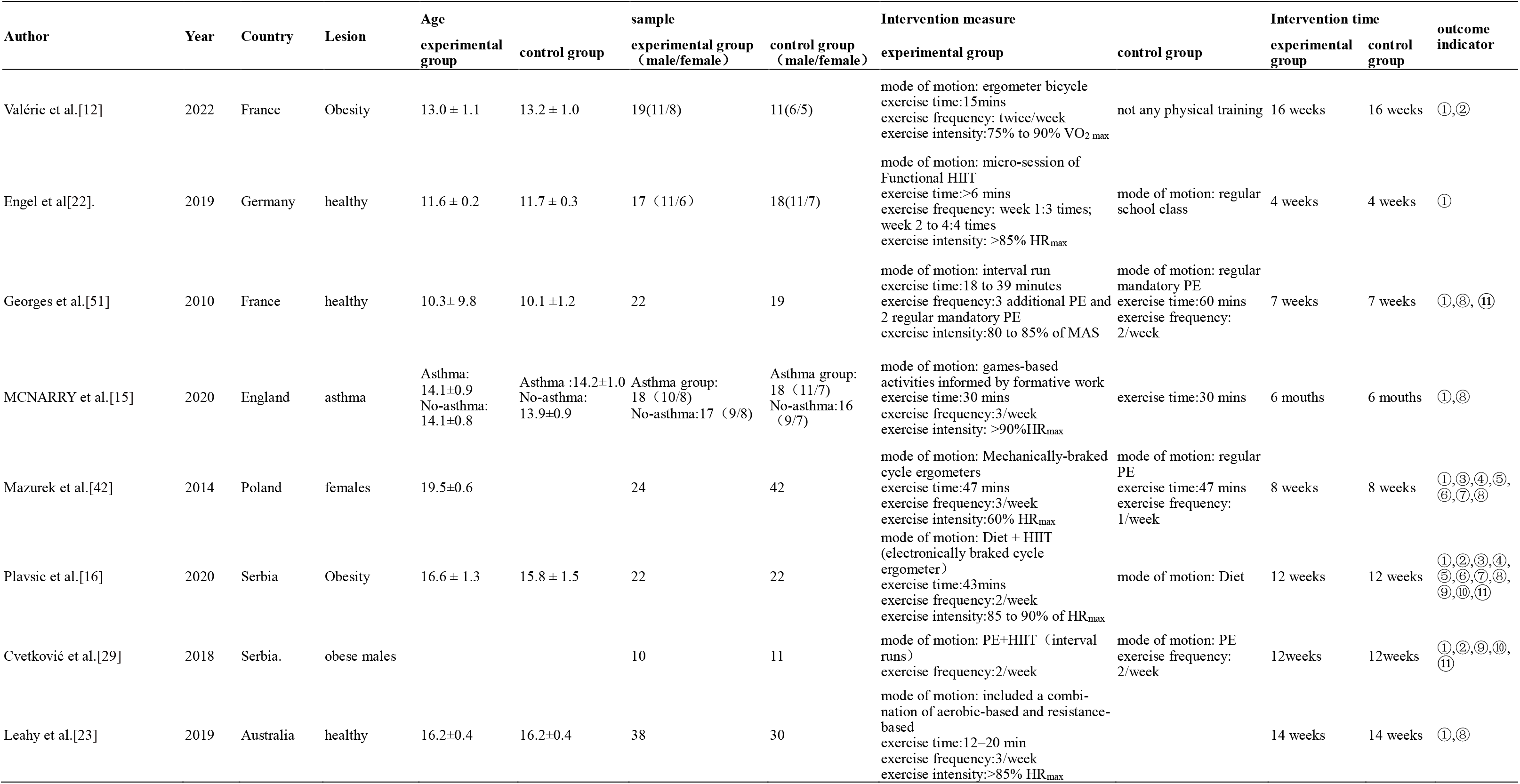
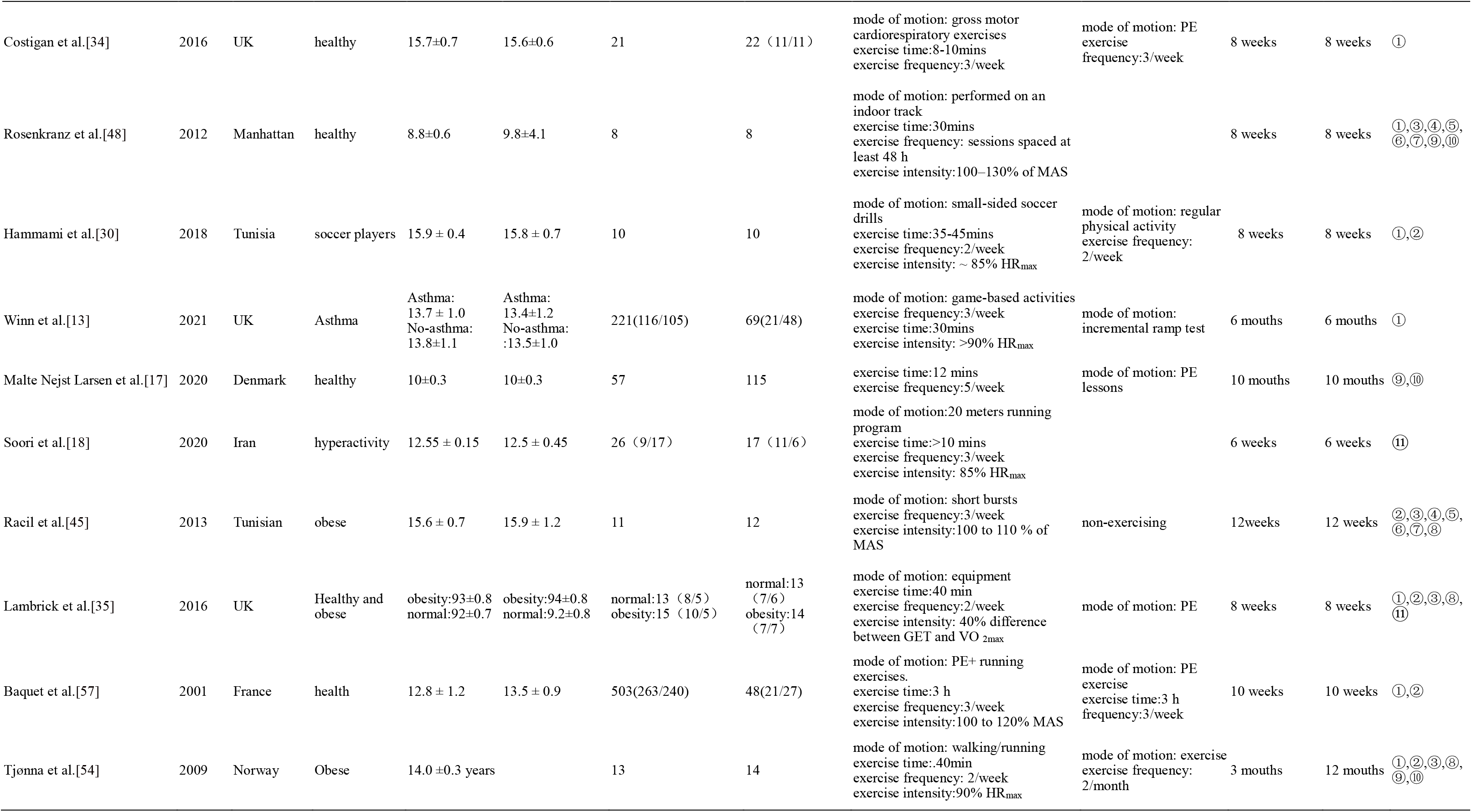
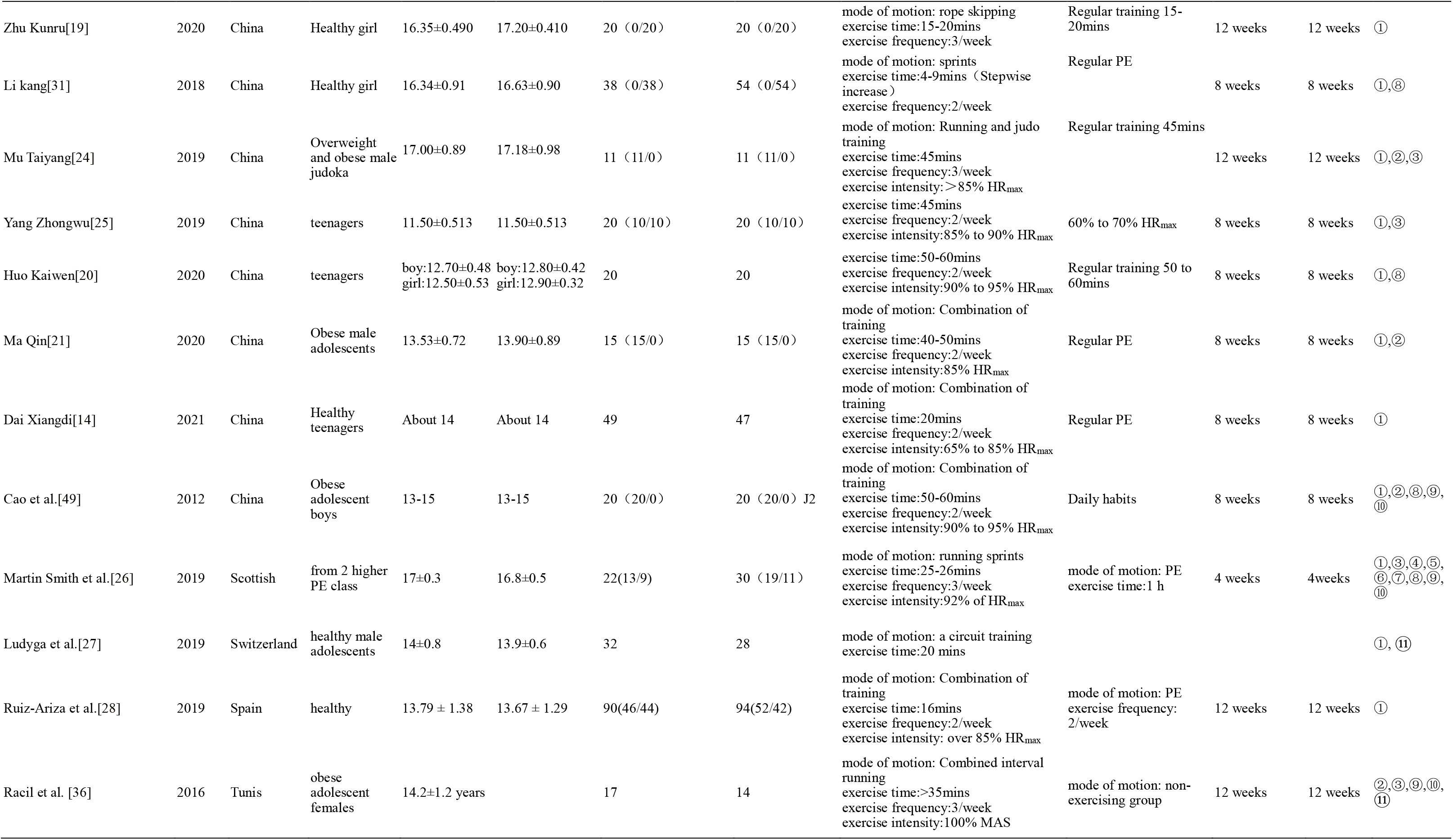
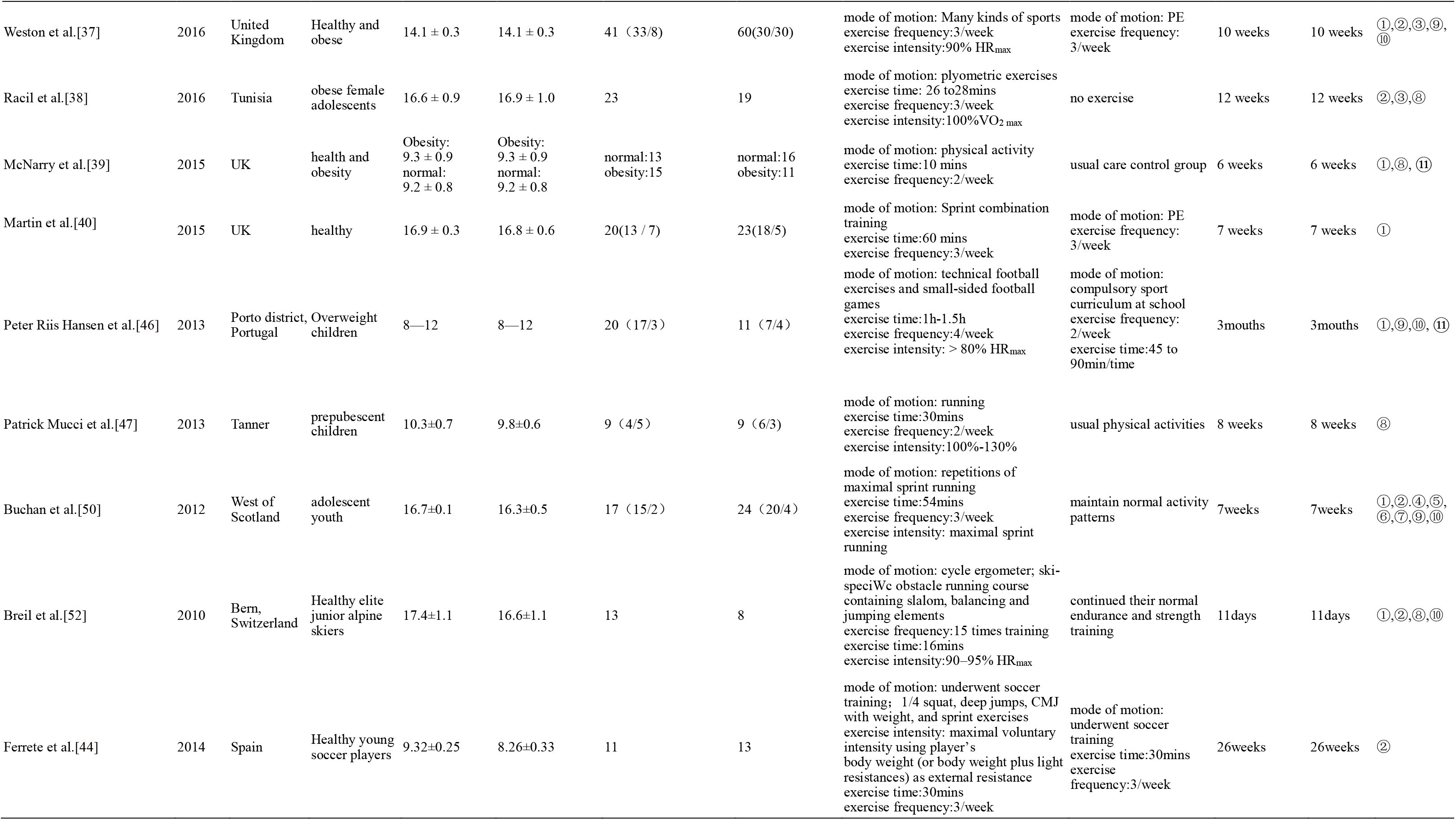
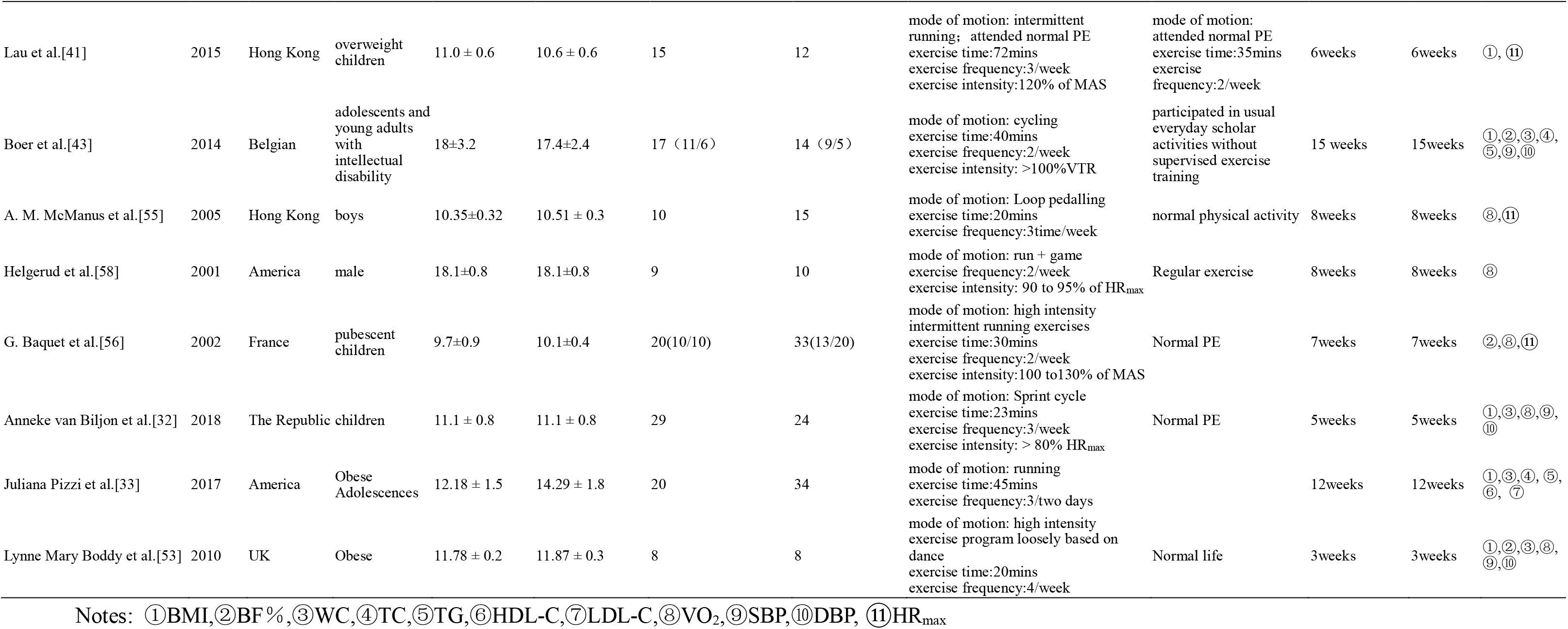
Basic features of the included studies

**Table 2.**
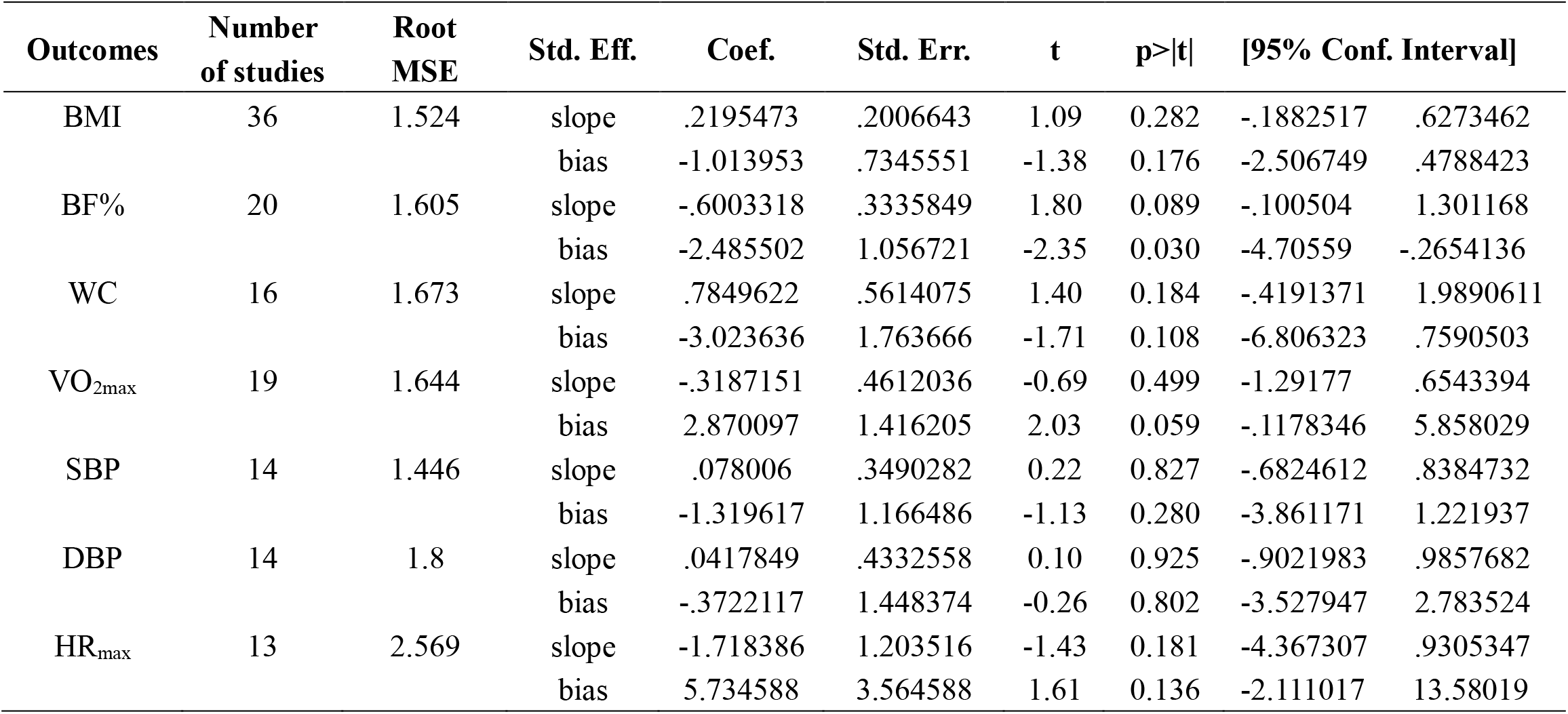
Three-line table of Egger’s Publishing Bias

### Study quality assessment (risk of bias) and sensitivity analysis

A total of 47 studies were included in this meta-analysis, with reasonable overall risk bias and good quality papers (Fig. 2). Egger’s examined BMI, BF%, WC, VO_2max_, SBP, DBP, and HR_max_ and found that BF% was at risk of publication bias (p<0.05) (Table 3). The sensitivity results showed that the overall data were stable (Supplemental Fig. 1-11).

**Fig. 2.**
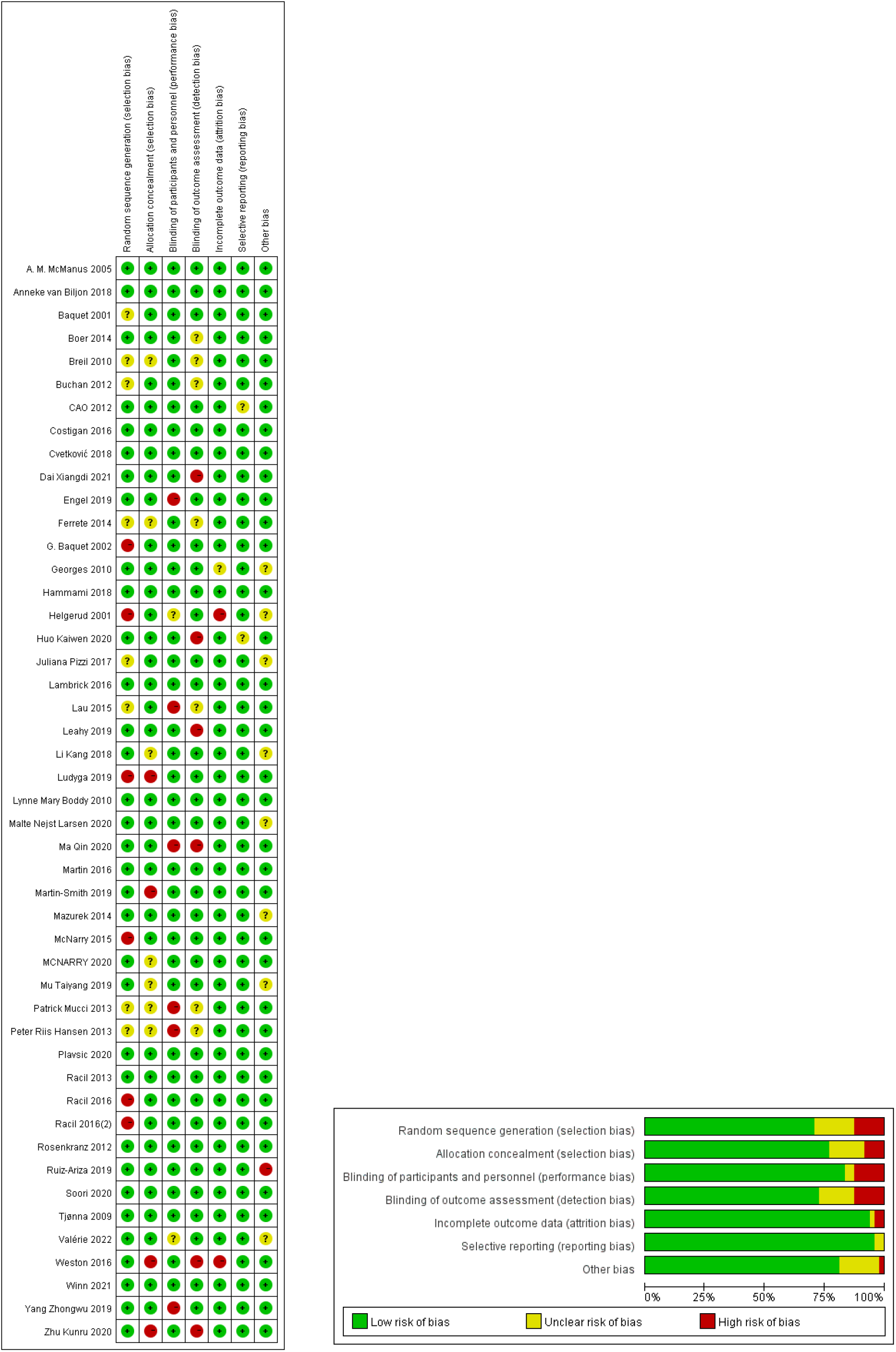
Analysis of the risk of bias according to the Cochrane Collaboration Guideline

**Table 3.**
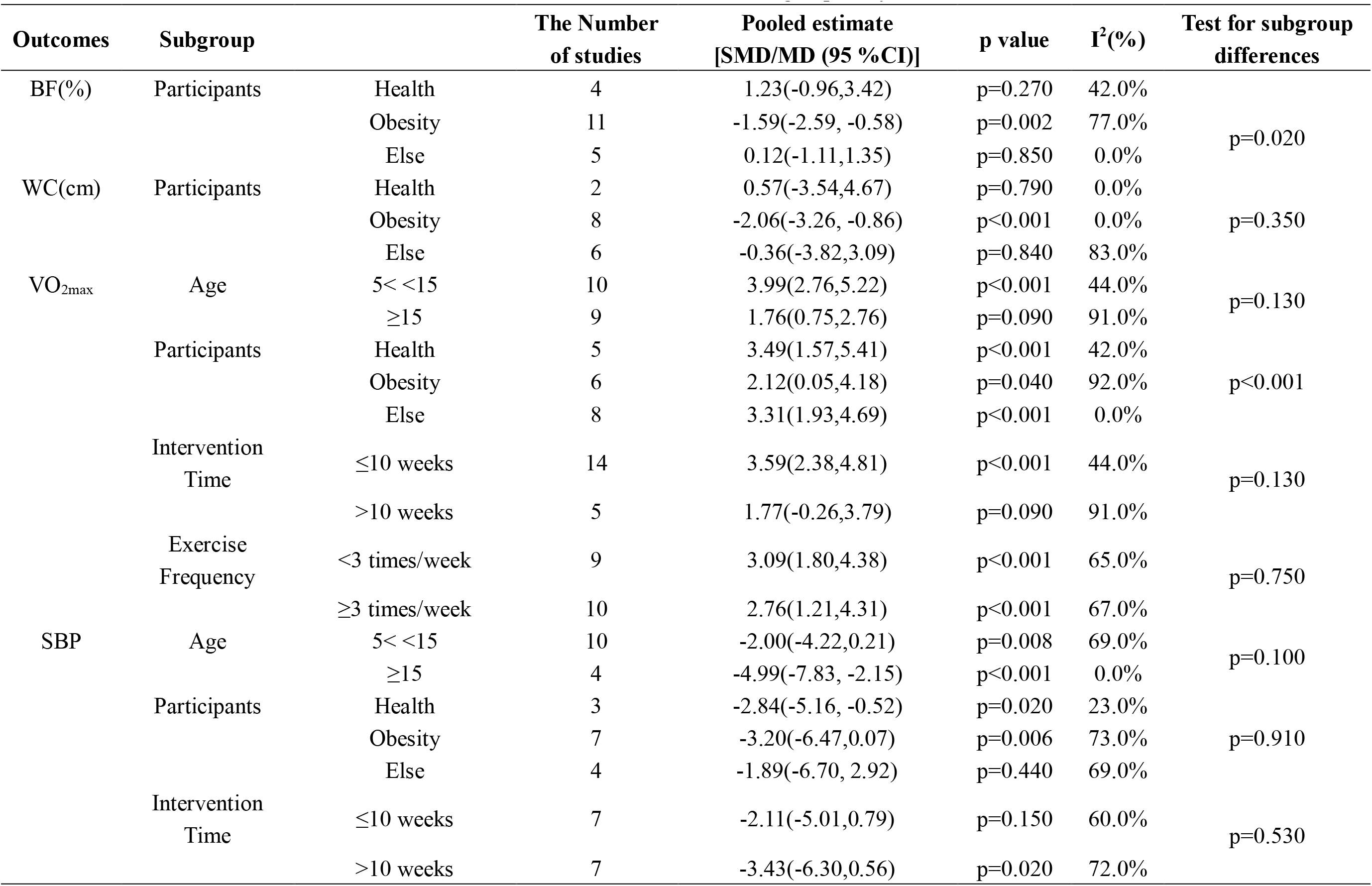
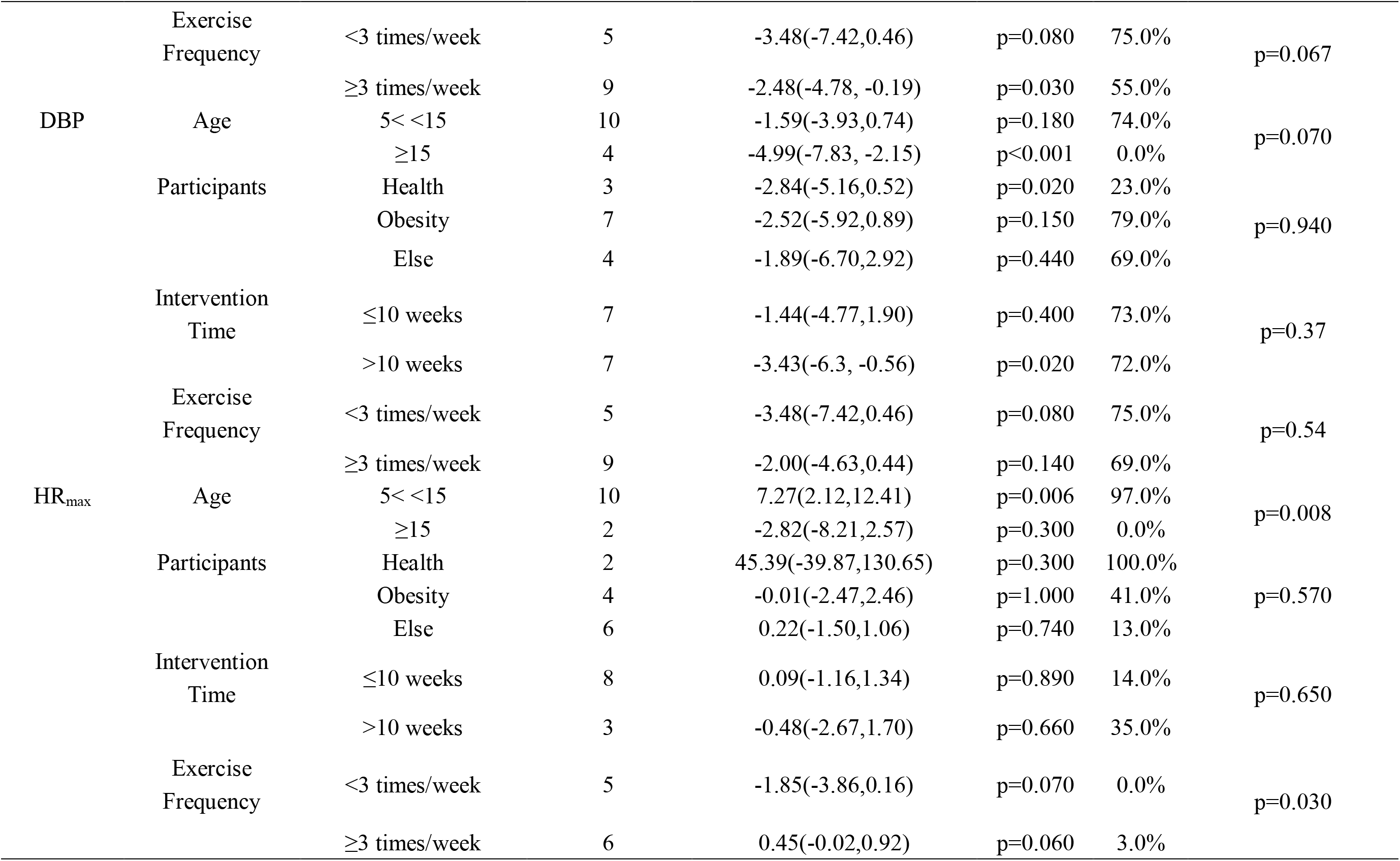
Three-line table of subgroup analysis

### Subgroup analysis

Substantial sources of heterogeneity can be explored through subgroup analyses. Due to differences in the age, participants, intervention time, and exercise frequency of HIIT intervention in children and adolescents included in the study, HIIT’s assessment of body morphology, CRF and cardiovascular disease metabolic risk indicators may be affected. Therefore, subgroup analyses have performed that were based on the age (5∼15 years old, ≥15 years old), participants (health, obesity, Else whose participants are not specified, not limited to healthy, obese people, etc.), intervention time (≤10 weeks, >10 weeks) and exercise frequency (≥3 times/week, <3 times/week), it was shown in Table 4.

### Meta-analysis results

#### Indicators of body morphology

In 42 studies, HIIT (n=1638) did not improve body morphology compared to the control group (n=1080).

In 37 studies, the HIIT group (n=1518) had no significant effect on BMI [MD=-0.30, 95% CI (−0.72,0.13), p=0.17] compared with the control group (n=954). In 20 studies, the HIIT group (n=829) had no significant effect on BF% [MD=-0.79,95% CI (−1.64,0.06), p=0.07] compared with the control group (n=394). In 16 studies, the HIIT group (n=314) had no significant effect on WC [MD=-1.24, 95% CI (−2.78,0.30) compared with the control group (n=359) (Fig. 3).

**Fig. 3.**
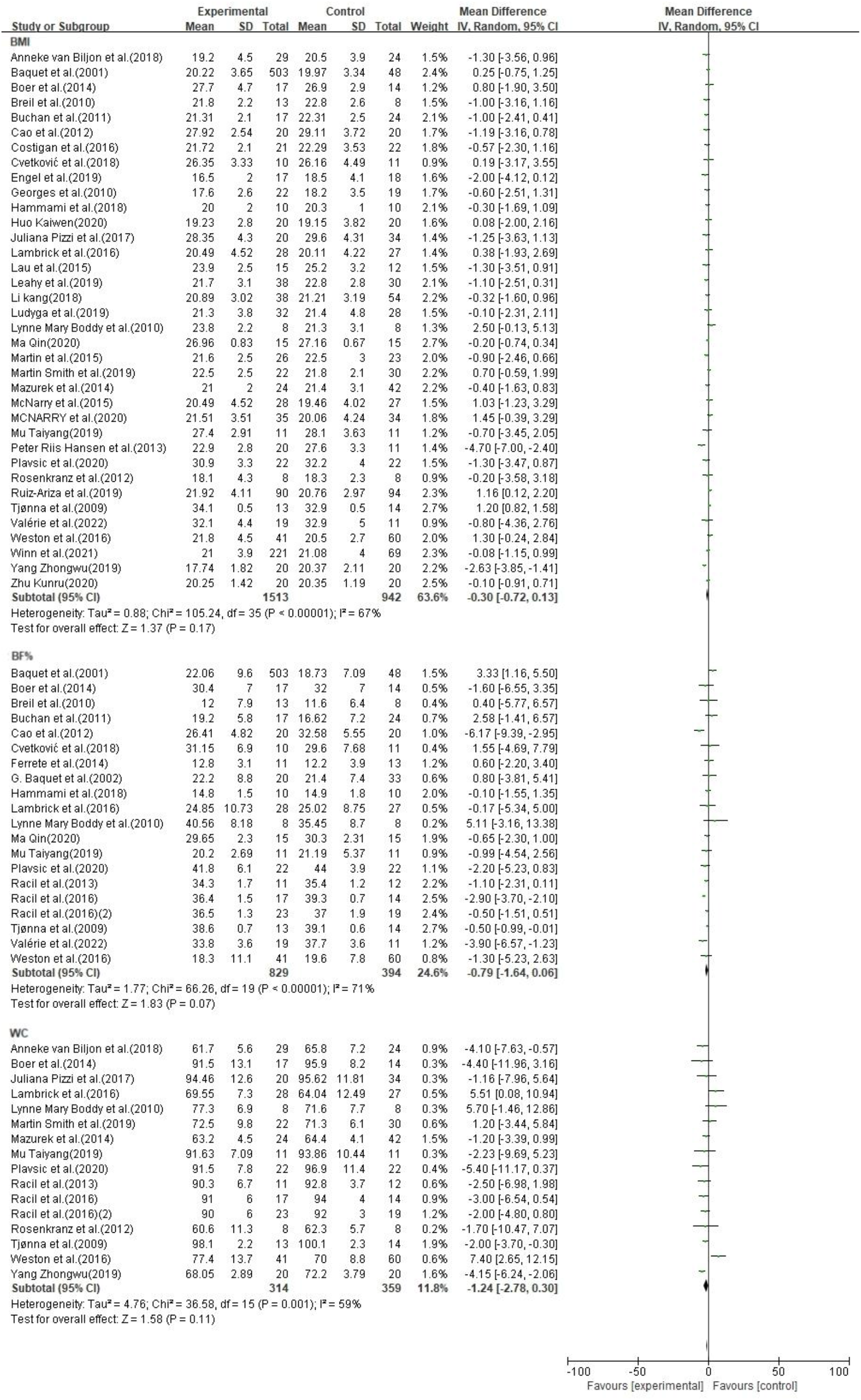
Forest plot of body morphology indicators

### CRF Indicators

In 32 studies, HIIT (n=702) effectively improved CRF indices compared with control groups (n=791), but clinical heterogeneity was high, so a subgroup analysis of CRF indexes was conducted (Fig. 4).

**Fig. 4.**
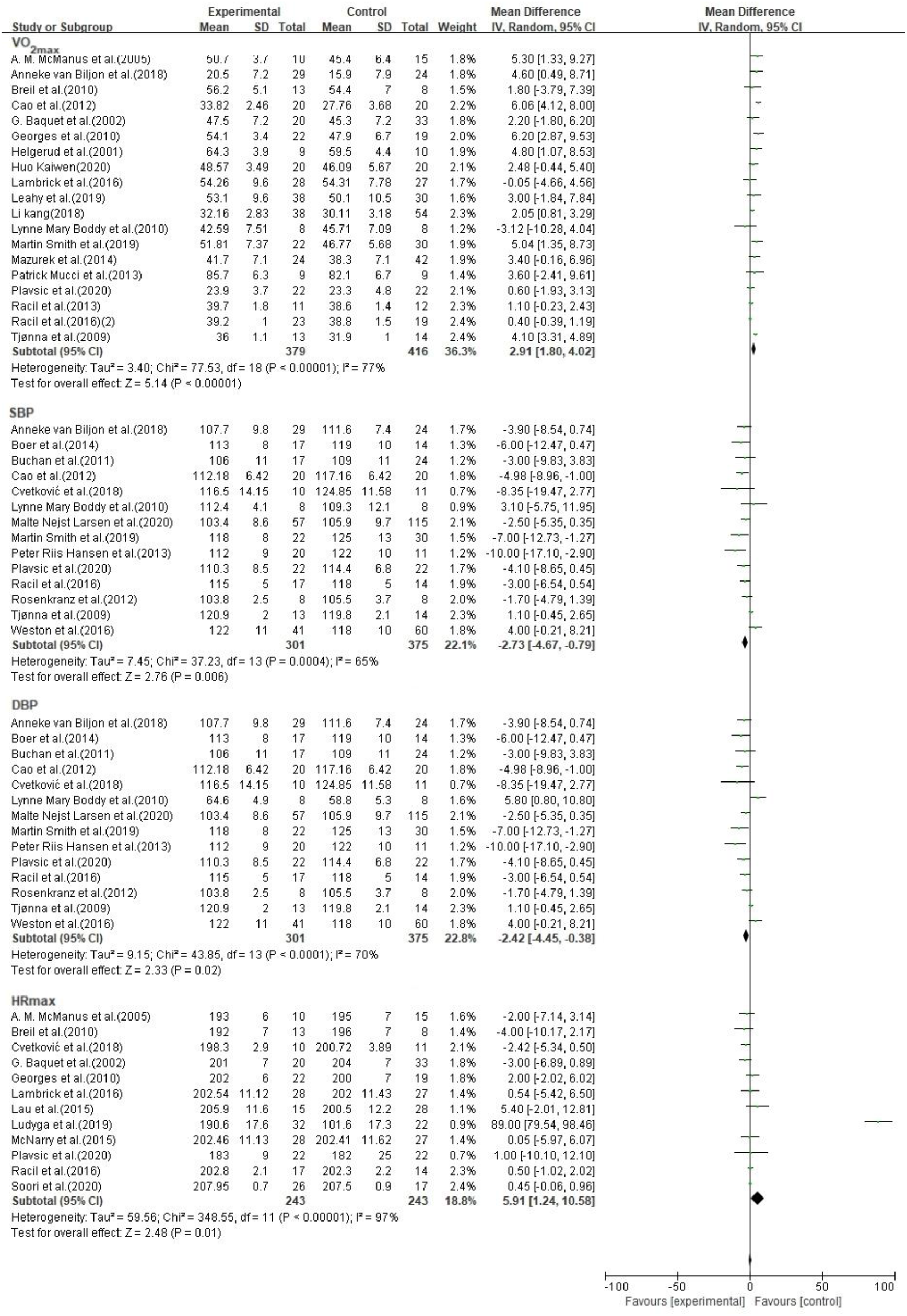
Forest plot of CRF indicators

In 19 studies, HIIT group (n=379) effectively increased VO_2max_ [MD=2.91, 95% CI (1.80, 4.02), p<0.001] compared with the control group (n=416) but with higher heterogeneity (I^2^=77%, p<0.001. The results of the subgroup analysis showed that HIIT was more effective in aged 5∼15 years (HIIT: n=179, control group: n=189), healthy children and adolescents (HIIT: n=133, control group: n=141), intervention time ≤10 weeks (HIIT: n=244, control group: n=292) and exercise frequency <3 times/week (HIIT: n=179, control group: n=209)

In 14 studies, HIIT group (n=301) effectively reduced SBP [MD=-2.73, 95% CI (−4.67, −0.79),p=0.006] compared with the control group (n=375) but with higher heterogeneity (I^2^=65%, p<0.001). The results of the subgroup analysis showed that HIIT was more effective in aged 5∼15 years (HIIT: n= 223, control group: n=285), else children and adolescents (HIIT: n=104, control group: n=122), intervention time ≤10 weeks (HIIT: n=145, control group: n=174) and exercise frequency ≥3times/week (HIIT: n= 219, control group: n=294).

In 14 studies, HIIT group (n=301) effectively reduced DBP [MD=-2.42, 95% CI (−4.45, −0.38), p=0.02] compared with control group (n=375) but with higher heterogeneity (I^2^=70%, p<0.001). The results of the subgroup analysis showed that HIIT was more effective in aged 5∼15 years (HIIT: n= 223, control group: n=285), else children and adolescents (HIIT: n=104, control group: n=122), intervention time ≤10 weeks (HIIT: n=145, control group: n=174) and exercise frequency ≥3 times/week (HIIT: n=219, control group: n=294).

In 12 studies, HIIT group (n=243) and the control group (n=233) had an effective increase in HR_max_ [MD=5.91, 95% CI (1.24, 10.58), p=0.01] compared with HIIT [MD=5.91, p=0.001] but with higher heterogeneity (I^2^=97%, p<0.001). The results of subgroup analysis showed that HIIT was more effective in aged 5∼15 years children (HIIT: n=208, control group: n=101), healthy children and adolescents (HIIT: n=95, control group: n=82), intervention time >10 weeks (HIIT: n=49, control group: n=47) and exercise frequency <3 times/week (HIIT: n=108, control group: n=120).

### Cardiovascular metabolic indicators

In 8 studies, compared with the control group (n=141), the HIIT group (n=186) significantly improved the metabolic risk index of cardiovascular disease, but some of the indicators were not statistically significant (Fig. 5).

**Fig. 5.**
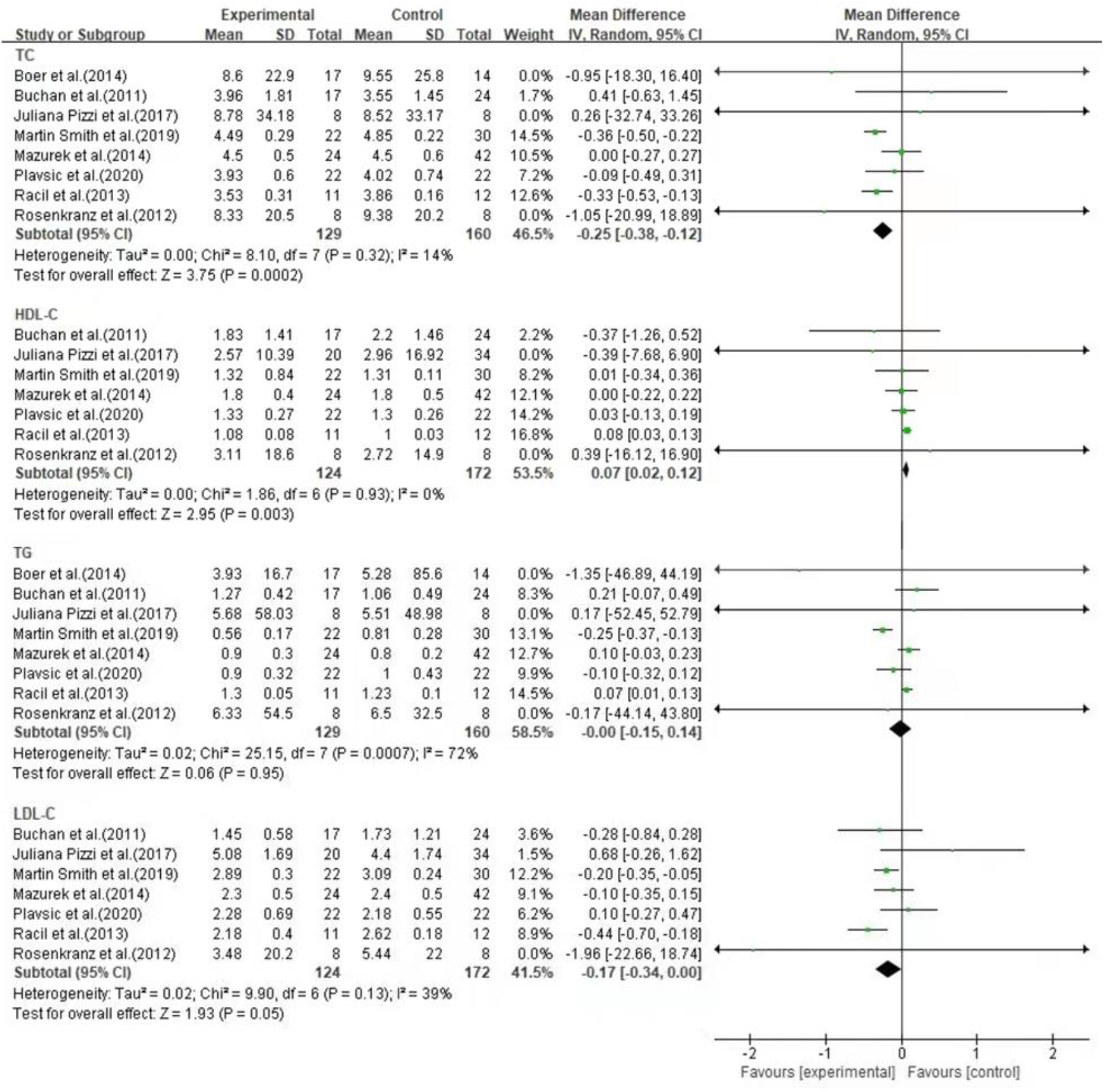
Forest plot of cardiovascular metabolic indicators

In 8 studies, HIIT (n=141) effectively reduced TC compared with control group (n=186) [MD=-0.27, 95%CI (−0.38, −0.17), p<0.001] with no significant heterogeneity (I^2^=14%, p=0.32). In 7 studies, compared with the control group (n=172), HIIT (n=124) effectively increased HDL-C [MD=0.07, 95% CI (0.02, 0.12), p=0.003] without significant heterogeneity (I^2^=0%, p=0.93). In 8 studies, the effect of HIIT on TG was not statistically significant between HIIT (n=141) and control group (n=186) [MD=-0.00, 95% CI (−0.15, 0.14), p=0.95]. In 7 studies, the effect of HIIT on LDL-C was not statistically significant between HIIT (n=124) and control group (n=172) [MD=-0.17, 95% CI (−0.34, 0.00), p=0.05].

## DISCUSSION

More and more studies have revealed that physical activity can significantly improve the physical health of children and adolescents[1]. Nevertheless, the global survey data shows that most children and adolescents do not meet the standards of physical activity guidelines, which seriously affects their current and future health[1]. To improve the level of physical activity of children and adolescents and achieve the goal of reducing the incidence of physical inactivity in children and adolescents by 15% in 2030[5], WHO issued the updated “2020 WHO Guidelines on Physical Activity and Sedentary Behavior” in 2020, which recommended that children and adolescents should engage in an average of not less than 60 minutes of moderate-to vigorous-intensity physical activity per day. Because children and adolescents are concentrated on campus most of the time, considering that physical activity is limited by time, HIIT is a good choice for its sports characteristics.

This study is the first to systematically evaluate the safety and efficacy of HIIT in terms of body shape, CRF, and cardiovascular disease metabolic risk indicators for children and adolescents of all ages (including health, obesity and disease) by synthesizing 47 eligible randomized controlled trials. With strict inclusion and exclusion criteria (age, exercise intensity, exercise frequency and exercise time), 47 studies were assessed. Available evidence shows that HIIT can significantly improve most indicators of children and adolescents’ health: improved CRF indexes (VO_2max_, SBP, DBP and HR_max_) and cardiovascular disease metabolic risk indicators (TG and HDL-C). In addition, there is insufficient evidence that HIIT improves body shape indicators (BMI, BF% and WC) and some cardiometabolic measures (TG, LDL-C) in children and adolescents.

Statistical heterogeneity consisted in most outcome measures due to multiple factors. First, our meta-analysis includes children and adolescents, who span a wide age range. Since this stage is pre-pubertal and adolescence, the developmental speed is related to age. Previous studies have shown that prepubertal children may obtain greater benefits in HIIT[7], and different developmental stages may affect the assessment of HIIT in body shape indicators. Secondly, the participants included in the study have healthy, overweight, obese, and partially diseases, together with children and adolescents with sports training experience, which led to differences in participants at baseline level and affected the assessment of HIIT outcomes. Third, the study intervention time ranges from 1.57 weeks to 40 weeks, and it is generally believed that a longer exercise intervention time is more likely to obtain greater benefits. Finally, the exercise frequency of participants is an essential part of exercise prescription or exercise program, and the exercise frequency of the included studies varied from 2 to 5 times/week, which may affect the evaluation of HIIT outcomes.

In addition, this study also determined the HIIT dose-response relationship: with interval running as the primary form of exercise, the exercise intensity is ≥80%VO_2max_/≥100%MAS/≥80%HR_max_. For healthy children and adolescents aged 5∼15 years old, the health benefit is the greatest when the intervention time is ≤10 weeks and the exercise frequency is 2∼5 times/week, while children and adolescents aged ≥15 years cannot recommend exercise doses due to the limited number of included studies.

### Effect of HIIT on body shape of children and adolescents

Previous studies have shown that HIIT can effectively improve the body shape indicator of children and adolescents. There is now insufficient evidence that HIIT improves body shape BMI, BF%, and WC indicators in children and adolescents. The findings are consistent with previous studies[7, 8], but there are diametrically opposite conclusions[9]. Although this study showed that HIIT had no significant effect on the body shape indicators of children and adolescents, it may have a greater impact on the reliability of the results due to the large range of subjects included in the study. Further analysis found that HIIT had a significant effect on obese children and adolescents with BF% [MD=-1.59, 95% CI (−2.59, −0.58), p=0.002] (Fig. 6) and WC [MD=-2.06, 95% CI (−3.26, −0.86]), p<0.001] (Fig. 7), the effect of the study results was positive, and it had no significant effect on BMI [MD=-0.91, 95% CI (−1.91, 0.09), p=0.08]. Metabolic disorders caused by overweight/obesity are the pathological basis of various metabolic diseases such as T_2_DM and CVD. Adverse metabolic phenotypes are highly associated with obesity in children and adolescents. Unfortunately, more than 50% of children and adolescents will carry obesity into adulthood, and the proportion increases with age[3]. In addition, children and adolescents with increased BMI will increase the risk of T_2_DM, stroke, coronary heart disease and cancer. As well as prospective cohort studies have shown that BF% and WC, the two body shape indicators, dropped to the normal range significantly reducing the prevalence of obesity-related diseases[59]. Therefore, a healthy body shape in children and adolescents is crucial.

**Fig. 6.**
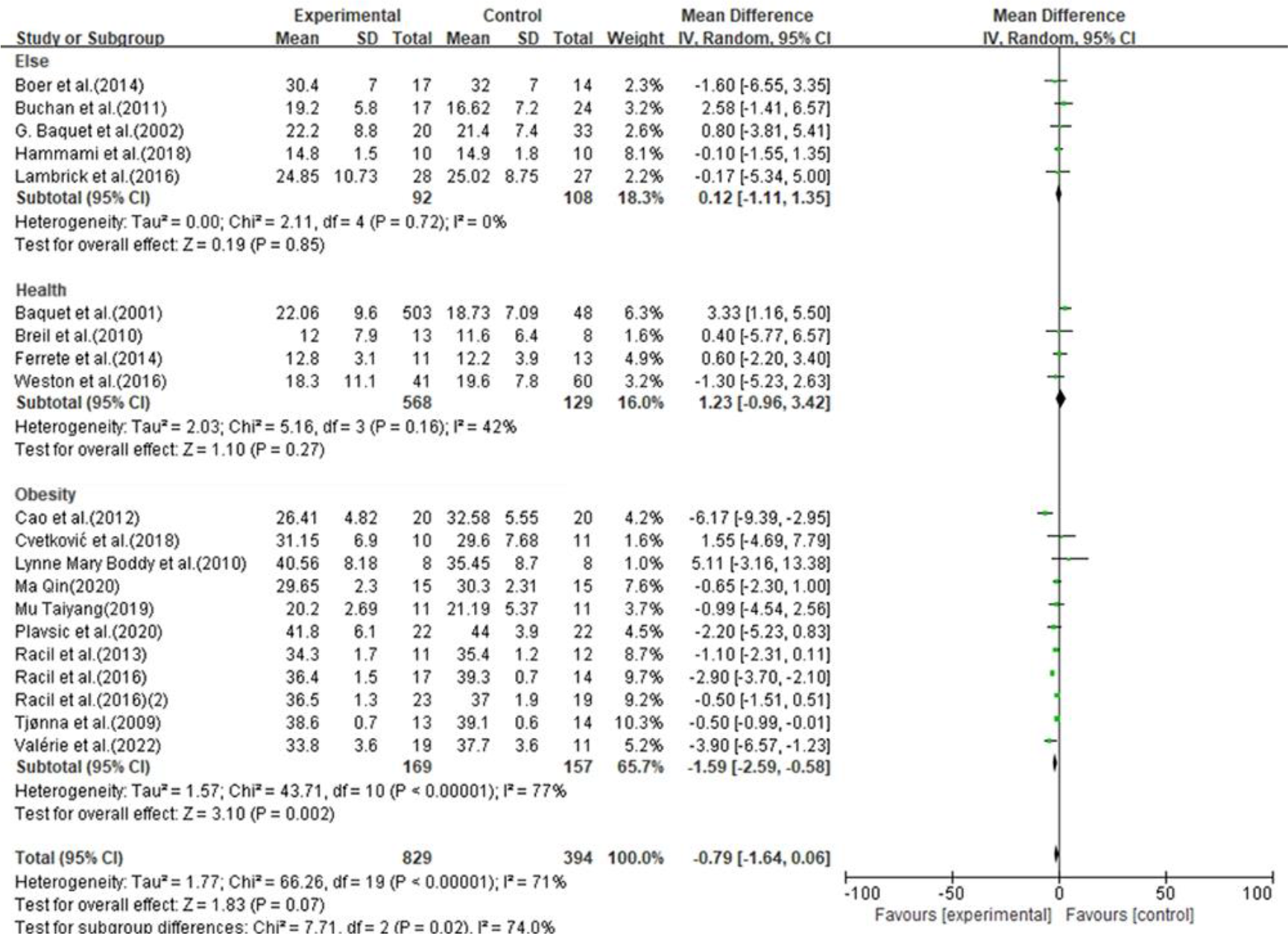
Participants subgroup analysis of BF% max in children and adolescents in the HIIT group and the control group

**Fig. 7.**
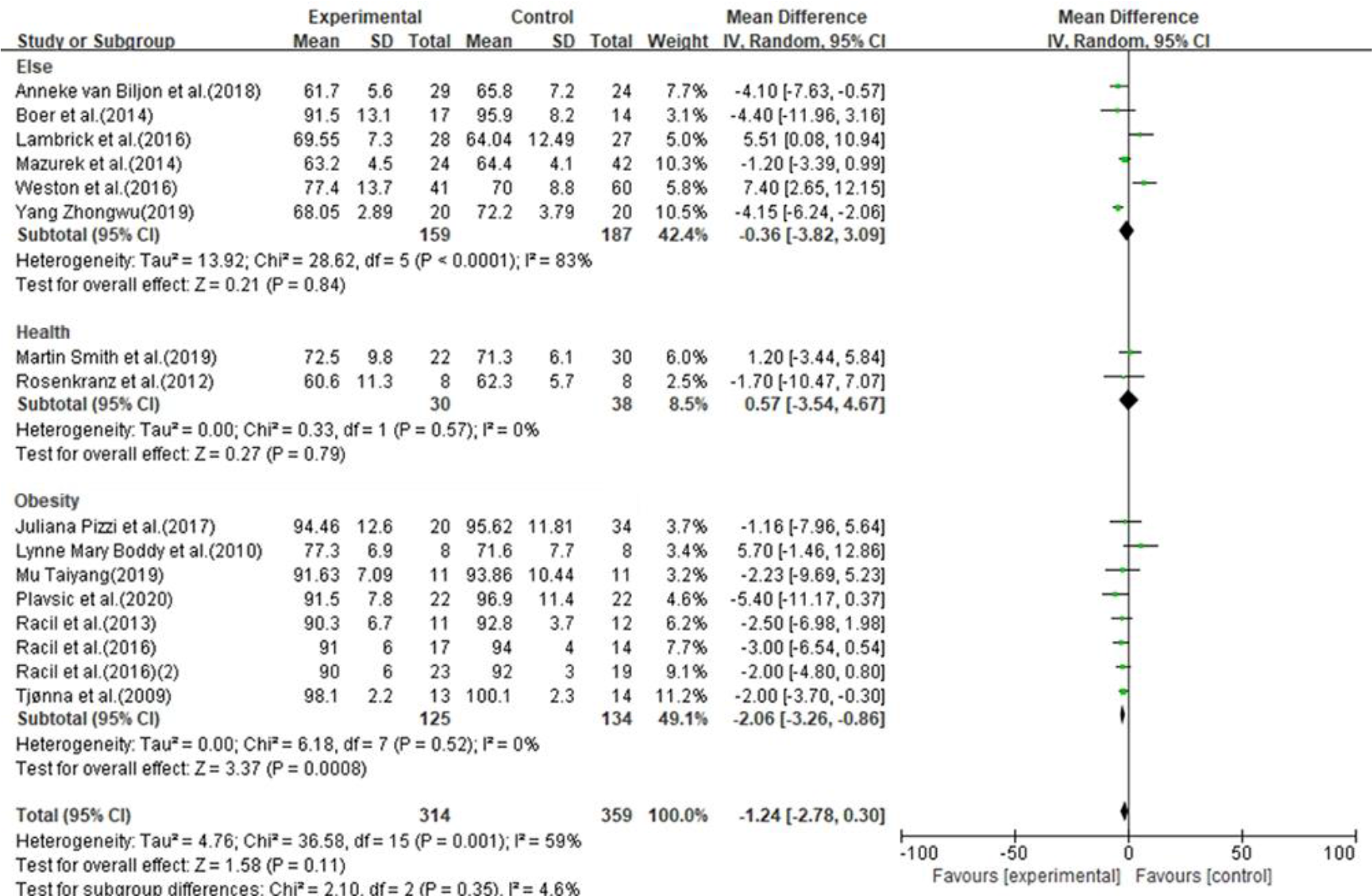
Participants subgroup analysis of WC max in children and adolescents in the HIIT group and the control group

At present, the key to the childhood obesity epidemic is inflammation and metabolic disorders. Anti-inflammatory and antioxidant interventions help regulate inflammation and metabolic disorders caused by obesity. Significant progress has been made in the prevention and treatment of obesity, but obesity-related diseases still cannot be effectively controlled. It is worth noting that majority of childhood and adolescent obesity is caused by insufficient physical activity. The prevalence of obesity in the physically inactive population has increased and there is still a potential risk in physically inactive non-obese individuals. However, it is gratifying that regular exercise can independently reduce the risks of obesity due to metabolic disorders, such as T_2_DM, CVD, and cancer, and can play a critical role in related diseases[4]. Particularly early exercise interventions are more effective in children and adolescents. Recent research has shown that running-based HIIT can improve the physical health of obese teenagers, and it is suggested to conduct a larger-scale survey of participants from different schools[10].

What’s more, we found a subset of athletes in healthy children and adolescents[30, 44, 52]. The research subjects were other children and adolescents with partial intellectual disability[43] and non-uniform subjects (the included participants were overweight, obese and healthy children and adolescents, and the data could not be classified and extracted)[35, 37]. There may be certain data risks due to the inconsistency of the population classification of the study subjects.

The sensitivity analysis of this meta-analysis results in high stability, and we can conclude that although HIIT does not improve body shape indicators in children and adolescents, its recommendation for obese children and adolescents should be retained.

### Effect of HIIT on CRF in children and adolescents

CRF has become one of the most extensive components of physical health research because of its strong correlation with health outcomes. Strong epidemiological evidence suggests that CRF is inversely associated with a high incidence of CVD, all-cause mortality and cancer in the healthy population. CRF can be used as a predictor of cancer mortality, and higher levels of CRF can independently reduce cancer mortality in women and men[60]. This study confirmed that HIIT is an effective method to improve CRF indicators in children and adolescents, and the findings are consistent with those of most meta-analysis studies. Because of the differences in whether research subjects are obese or not, there is still controversy about HIIT replacing MICT[9], still, it has gradually been widely accepted because HIIT is more cost-effective. Although the relationship between physical activity and the incidence and mortality of CVD, T_2_DM and cancer, etc. and its preventive effect have been demonstrated, children and adolescents still face the plight of insufficient physical activity. Extensive research has been carried out in the field of sports medicine, and a great many of research results has been obtained.

Our meta-analysis results show that HIIT can effectively improve the CRF index, but there is some statistical heterogeneity. We conducted a subgroup analysis of CRF index heterogeneity, and found that the participants were the source of VO_2max_ heterogeneity; age and exercise frequency are the source of HR_max_ heterogeneity. However, age, participants, intervention time and exercise frequency are not heterogeneous sources of SBP and DBP.

VO_2max_ is the gold standard for evaluating CRF. Obese children and adolescents exhibit considerable limitations in physical activity due to skeletal muscle oxidative disorders in obese children and adolescents. Considering the influence of weight on VO_2max_ level, The VO_2max_ of obese children and adolescents’ overall baseline level is low, and its change is more sensitive to HIIT intervention. In contrast, the VO_2max_ level in healthy children and adolescents tends to be stable.

Our results show that age is the main source of heterogeneity in HR_max_. According to the actual situation of included studies, in this subgroup analysis, ages between 5∼15 years old and ≥15 years old were divided into groups. Although not grouped by pre-pubertal and adolescence, the actual grouping covers prepubertal and adolescent ages to a greater extent.

In this subgroup analysis, the proportion of obese adolescents aged ≥15 years in this subgroup analysis is more[16, 36, 38, 49]. Children are more resilient, making them more tolerant of HIIT and more resistant to fatigue than adolescents. More importantly, HIIT is more closely related to children’s exercise habits, combined with children’s exercise patterns (game)[35]and reward mechanisms[53, 55, 56]in the included studies, which make pre-adolescent children more autonomous and motivated to participate.

It is worth noting that high-intensity training triggers autonomic nervous disturbances and high exercise frequency triggers fatigue accumulation while also causing a “compensatory effect”. Subgroup analysis included 6 studies [18, 36, 41, 51, 52, 55] with exercise frequency ≥3 times/week. Since children’s tolerance, autonomy and motivation to participate are more advantageous than adolescents, a study has shown that HR_max_ has the potential to predict VO_2max_, which also further confirms the role of age in HR_max_ and VO_2max_ [MD = 3.99, 95% CI (2.76, 5.22), p<0.001]. Future research is supposed to pay attention to the dose-response relationship in exercise frequency towards CRF and focus on the interest and reward mechanism of HIIT in pre-puberty children during the specific implementation process. Simultaneously, improve the design of HIIT in adolescents to enhance their initiative and motivation to participate.

According to the available evidence, we can conclude that the intervention time ≤10 weeks, the frequency of exercise <3 times/week, and healthy children and adolescents aged 5∼15 are more sensitive to improving VO_2max_ after HIIT intervention. The intervention time >10 weeks, the frequency of exercise <3 times/week, and healthy children and adolescents aged 5∼15 are more sensitive to improving HR_max_ after HIIT intervention. Although partial results fail to explain the source of heterogeneity, VO_2max_ and HR_max_, which are considered to the most important outcome indicators for evaluating CRF, well explain the heterogeneity. Despite certain sources of heterogeneity in SBP and DBP, their sensitivity analysis still suggests that the results are stable due to their relatively mature measurement methods. In summary, HIIT is recommended for improving CRF indicators in children and adolescents.

### Effect of HIIT on metabolic risk indicators of CVD in children and adolescents

At present, insufficient physical activity, unreasonable dietary structure, obesity, metabolic syndrome and other metabolic risk factors for cardiovascular disease are gradually superimposed, resulting in a sharp increase in the risk of CVD and T_2_DM. Metabolic risk factors of cardiovascular disease in children and adolescents during this period have a significant influence on the onset of adulthood[3]. The good news is that cardiovascular risk factors are largely preventable, especially the effectiveness of exercise in improving metabolic risk markers for cardiovascular diseases is supported by substantial evidence.

According to our research, a meta-analysis of HIIT intervention in children and adolescents, HIIT was effective in improving TC and HDL-C that are metabolic risk index of cardiovascular disease in children and adolescents, still, the effect on TG and LDL-C appeared to be insignificant. The results of this meta-analysis are similar to the previous meta-analysis results[7], but there are some differences in the analysis of HIIT evaluation on children and adolescents in another study[8], whose difference may be caused by HIIT’s limited improvement of healthy children and adolescent blood lipids[42]. Most of the crowds we have included are healthy children and adolescents, and their incorporations are mostly obese children and adolescents.

Our convergence analysis results show that HIIT can effectively improve the level of TC and HDL-C, without significant heterogeneity. Racil[45] and Boer et al.[43] reported that HIIT reduced TC in children and adolescents with a clinically significant (P<0.05) and low risk of bias assessment, which are encouraging findings. Notably, Racil et al.[45] evaluated obese girls and Boer et al.[43] evaluated children and adolescents with disabilities. Racil[45] and Juliana et al.[33] reported that HIIT reduced HDL-C in children and adolescents with clinical significance (p<0.05).Racil et al.[45] and Plavsic et al.[16] evaluated objects are obese girls. The limitations of study methods and subjects for TG and HDL-C in the included studies may confuse the results, so caution should be exercised in interpreting these results. Though HIIT does not improve TG and LDL-C levels, the importance of TG as an independent risk factor for CVD cannot be ignored. The effect of HIIT on LDL-C [MD=-0.17, 95% CI (−0.34, 0.00), p=0.05] was at a statistical critical value. After excluding the included studies one by one, it found that the study by Juliana et al. [33] had a high risk. After the exclusion, the effect of HIIT on LDL-C was statistically significant (p<0.001) and there was no significant heterogeneity (I^2^=24%, p=0.25).

The sensitivity analysis results in this meta-analysis were highly stable and showed no significant heterogeneity. We can conclude that HIIT is effective in improving TC and HDL-C in children and adolescents, with little effect on TG and LDL-C. Considering the small sample size included in this meta-analysis, future research requires expanding the sample size further.

### Assessment of diet and leisure-time physical activity

35 studies[13-15, 17, 19-35, 39, 41, 43, 47-53, 55-58] did not describe dietary assessment in detail. 12studies[12, 16, 19, 36–38, 40, 42, 44–46, 54]informed not to change dietary habits. 35 studies[12-15, 17,18, 20-28, 30-35, 39, 41, 43, 45, 48-53, 55-58]did not describe leisure-time physical activity in detail, and12studies[16, 19, 29, 36–38, 40, 42, 44, 46, 47, 54]conducted detailed assessments of leisure-time physical activity. Although 74.47% of the studies did not describe the diet in detail, it can be understood as maintaining the original eating habits and physical activities by reading the full text. Furthermore, although 25.53% were assessed for dietary habits and leisure-time physical activity, the impact of dietary and leisure-time physical activity assessments on outcomes was not elucidated. It is worth noting that maintaining the original dietary habits and physical activity helps to rule out the influence of diet on HIIT outcomes, simultaneously diet and leisure physical activity may be important limiting factors and sources of bias.

### Adverse events and compliance

In this meta-analysis, although only 21 studies [12, 16, 18, 22, 26–29, 37, 38, 40, 43–45, 48, 50–52, 54, 55, 58] (42.55%) reported withdrawal events due to family and subjective will (193 people dropped out, accounting for 6.445 of the total study, with a dependency of 93.56%). In addition, there were sicked children and adolescents in the included studies[13, 15, 18, 43], but no adverse events occurred, and the safety and dependence were good. A previous meta-analysis of HIIT reported the occurrence of adverse events such as leg discomfort, joint sprains, asthma, myocardial infarction, etc[61]. The occurrence of adverse events was attributed to the study subjects belonging to high-risk groups for adverse events, such as patients with coronary heart disease and hypertension, but there seemed to be no difference in adverse events between the HIIT group and the control group. Besides, medical supervision was described in 55.32% of the included articles in this meta-analysis. Therefore, these factors may cause biases, but the results were skewed towards more positive effects.

### Advantages and limitations

Advantages of this study: (ⅰ) Retrieval was not limited by publication date. (ⅱ) Research participants were not limited to specific children and adolescents but includes all children and adolescents, regardless of health, disease, etc. (ⅲ) Subgroup analysis was carried out to explain the heterogeneity of research results, especially the analysis of exercise dose variables such as age (prepubertal and adolescence), research subjects, intervention time and exercise frequency, which were often ignored by previous studies. (ⅳ) No-blank control was excluded in this study, and unbalanced results caused due to non-blank control were avoided.

Limitations of this study: (ⅰ) Although this review strictly implemented the retrieval strategy, due to limited conditions, only the literature published in Chinese and English were retrieved, and there may still be some publication bias due to the lack of a small number of published literatures. (ⅱ) This study only included the information on children and adolescents in school, but lacks data about children and adolescents outside of school, which may have a certain influence on the conclusion. (ⅲ) Quality of included studies may be another factor, 12.77% of the studies did not use the randomized control model, and 17.02% of the studies reported the randomized control model, but did not describe the randomized process. (ⅳ) Although guidelines for HIIT have been established, the details of some guidelines still need to be refined. (ⅴ) The biggest limiting factor may be that the age span of the individuals included in the study was large, and important influencing factors such as exercise intensity, frequency, and time were not completely consistent, and the heterogeneity was considerable.

Although subgroup analyses have been performed, some results still could not clarify the source of heterogeneity. For example, Sexual differences in developmental rates during childhood and adolescence (due to the mixed-gender or too small sample size for subgroup analysis), different methods of outcome measures, and diet and leisure-time physical activity may all be sources of heterogeneity. In addition, the included studies did not disaggregate by gender, and the impact of gender on children and adolescents was still unclear, so gender differences should be fully considered in future research.

## CONCLUSION

HIIT is safe, effective and less time-consuming for child and adolescent health. Because of its potential to improve body shape, CRF, and cardiovascular disease risk markers, it should be incorporated into the daily management of physical activity in children and adolescents. More importantly, the effect of HIIT has a higher consistency in gender, population, and age (pre-adolescence and adolescence), so it has a higher generality in improving physical health. Although there were dropouts and losses to follow-up during this process, no adverse events caused by HIIT occurred. These findings highlight the potential role of HIIT as a strategy for improving the health between children and adolescents. Considering the lack of more detailed standards for HIIT interventions in the included studies, it is worth studying specific HIIT interventions (optimal exercise interval time and interval intensity) in different ages, genders and participants to make HIIT more effective and scientific.

In conclusion, strengthening medical supervision and adequate warm-up before exercise is more feasible for the promotion of HIIT in children and adolescents.

## Data Availability

All data produced in the present work are contained in the manuscript.

## Acknowledgment

The completion of the thesis is very grateful to the co-funding of the Teaching Reform and Innovation Project of Shanxi Provincial Department of Education (J2021967) and the Teaching Reform and Innovation Project of Fenyang College of Shanxi Medical University (FJ202013). Many thanks to the authors for their hard work.

## Author’s contribution

Menjie made the agreement on this manuscript and revised it; Ma Jia, Zou Shuangling and Xiang Chenmin run a database search, selected included meta-analysis, evaluate its quality and extract data; Xiang Chenmin, Zou Shuangling and Wang Junli made statistical analysis; Li Shufeng contributed to the formulation of the protocol and revised the manuscript. All authors have read and approved the final version of the manuscript and agreed to the author’s presentation order.

