## Supplementary material for "High-Intensity Interval Training Improves physical morphology, Cardiopulmonary Fitness and Metabolic Risk Indicators of Cardiovascular Disease in Children and Adolescents: A Systematic Review and Meta-Analysis": Supplementary Material.docx

**Supplemental Tables:**

**File 1** Search Strategy.

**Pubmed:**

| **Items** | **Search Terms** | **Search Results** |
| --- | --- | --- |
| #1 | Adolescent [MeSH Terms] | 2148189 |
| #2 | (((((Adolescence [Title/Abstract]) OR (Teens [Title/Abstract])) OR (Teenagers [Title/Abstract])) OR (Youths [Title/Abstract])) OR (Female Adolescents [Title/Abstract])) OR (Male Adolescents [Title/Abstract]) | 113579 |
| #3 | #1 or #2 | 2175701 |
| #4 | High-Intensity Interval Training [MeSH Terms] | 1636 |
| #5 | ((((High-Intensity Interval [Title/Abstract]) OR (High-Intensity Intermittent [Title/Abstract])) OR (High-Intensity Intermittent Exercises [Title/Abstract])) OR (Sprint Interval Trainings [Title/Abstract])) OR (HIIT[Title/Abstract]) | 3458 |
| #6 | #4 or #5 | 3849 |
| #7 | randomized controlled trial [MeSH Terms] | 155174 |
| #8 | (((RCT[Title/Abstract]) OR (Randomized [Title/Abstract])) OR (Randomized Clinical [Title/Abstract])) OR (Controlled Clinical Trials [Title/Abstract]) | 611902 |
| #9 | #7 or #8 | 694148 |
| #10 | #9 AND #6 AND #3 | 92 |

**Cochrane Library:**

| **Items** | **Search Terms** | **Search Results** |
| --- | --- | --- |
| #1 | MeSH descriptor: [Adolescent] explode all trees | 108630 |
| #2 | (Adolescence): ti,ab,kw OR (Teens):ti,ab,kw OR (Teenagers):ti,ab,kw OR (Youths):ti,ab,kw | 8461 |
| #3 | #1 OR #2 | 114958 |
| #4 | MeSH descriptor: [High-Intensity Interval Training] explode all trees | 588 |
| #5 | (High-Intensity Interval): ti,ab,kw OR (High-Intensity Intermittent):ti,ab,kw OR (High-Intensity Intermittent Exercises):ti,ab,kw OR (Sprint Interval Trainings):ti,ab,kw OR (HIIT):ti,ab,kw | 3328 |
| #6 | #4 OR #5 | 3328 |
| #7 | MeSH descriptor: [Randomized Controlled Trial] explode all trees | 119 |
| #8 | (RCT): ti,ab,kw OR (Randomized):ti,ab,kw OR (Randomized Clinical):ti,ab,kw OR (Controlled Clinical Trials):ti,ab,kw | 1002643 |
| #9 | #5 or #6 | 1002643 |
| #10 | #7 and #8 and #9 | 143 |

**Embase**:

| **Items** | **Search Terms** | **Search Results** |
| --- | --- | --- |
| #1 | 'adolescent'/exp OR 'adolescent' OR 'adolescence'/exp OR 'adolescence' OR 'teens' OR 'teenagers' OR 'youths' OR 'female adolescents' OR 'male adolescents' | 1971546 |
| #2 | 'high-intensity interval training'/exp OR 'high-intensity interval training' OR 'high-intensity interval' OR 'high-intensity intermittent' OR 'high-intensity intermittent exercises' OR 'sprint interval trainings' OR 'hiit'/exp OR 'hiit' | 5171 |
| #3 | 'randomized controlled trial' OR 'rct' OR 'randomized' OR 'randomized clinical' OR 'controlled clinical trials' | 1276675 |
| #4 |  | 151 |

**Web of science:**

| **Items** | **Search Terms** | **Search Results** |
| --- | --- | --- |
| #1(participents) | TS=(adolescent OR Adolescence OR Teens OR Teenagers OR Youths OR Female Adolescents OR Male Adolescents) | 2837751 |
| #2(intervention) | TS=(High-Intensity Interval Training OR High-Intensity Interval OR High-Intensity Intermittent OR High-Intensity Intermittent Exercises OR Sprint Interval Trainings OR hiit) | 5171 |
| #3(study) | TS=(randomized controlled trial OR RCT OR Randomized OR Randomized Clinical OR Controlled Clinical Trials) | 1435301 |
| #4 | #1 AND #2 AND #3 | 345 |

**Science Direct (2015-now):**

| **Search Terms** | **Search Results** |
| --- | --- |
| (Adolescent OR Teens OR Youths) AND (High-Intensity Interval Training OR High-Intensity Intermittent Exercises OR Sprint Interval Trainings) AND (randomized controlled trial OR RCT OR Controlled Clinical Trials) | 2948 |

**CNKI：**

| **Search Terms** | **Search Results** |
| --- | --- |
| (SU=''青少年" OR SU="儿童'' OR SU="女青年" OR SU="男青年" OR SU="男儿童" OR SU="女儿童" OR SU="男运动员" OR SU="女运动员") AND (SU="高强度间歇训练" OR SU="冲刺间歇训练" OR SU="高强度间歇跳绳训练" OR SU="间歇训练" OR SU="有氧间歇训练" OR SU="无氧间歇训练" OR SU="长间歇训练" OR SU="短间歇训练" OR SU="高强度有氧间歇训练") AND (SU="随机对照试验" OR SU="随机化" OR SU="随机分配" OR SU="单盲" OR SU="双盲" OR SU="对照" OR SU="对照试验") | 40 |

**Wanfang：**

| **Search Terms** | **Search Results** |
| --- | --- |
| (主题:"青少年" or 主题:"儿童" or 主题:"女青年" or 主题:"男青年" or 主题:"男儿童" or 主题:"女儿童" or 主题:"男运动员" or 主题:"女运动员") and (主题:"高强度间歇训练" or 主题:"冲刺间歇训练" or 主题:"高强度间歇跳绳训练" or 主题:"间歇训练" or 主题:"有氧间歇训练" or 主题:"无氧间歇训练" or 主题:"长间歇训练" or 主题:"短间歇训练" or 主题:"高强度有氧间歇训练") and (主题:"双盲" or 主题:"单盲" or 主题:"对照" or 主题:"随机化" or 主题:"随机对照试验" or 主题:"随机分配" or 主题:"对照实验") | 15 |

**VIP：**

| **Search Terms** | **Search Results** |
| --- | --- |
| (M=青少年 OR M=儿童 OR M=女青年 OR M=男青年 OR M=男儿童 OR M=女儿童 OR M=男运动员 OR M=女运动员) AND (M=高强度间歇训练 OR M=冲刺间歇训练 OR M=高强度间歇跳绳训练 OR M=间歇训练 OR M=有氧间歇训练 OR M=无氧间歇训练 OR M=长间歇训练 OR M=短间歇训练 OR M=高强度有氧间歇训练) AND (M=随机对照试验 OR M=随机化 OR M=随机分配 OR M=单盲 OR M=双盲 OR M=对照 OR M=对照实验) | 0 |

**File 2:** Excluded Studies and Reason for Exclusion.

| **N** | **First Author（Year）** | **Title** | **Reason for exclusion** |
| --- | --- | --- | --- |
| 1 | T. Matsuo (2015) [1] | Effect of aerobic exercise training followed by a low-calorie diet on metabolic syndrome risk factors in men | Age out of range |
| 2 | G. Baquet (2001) [2] | High-Intensity Aerobic Training During a 10 Week One-Hour Physical Education Cycle: Effects on Physical Fitness of Adolescents Aged 11 to 16 | No result measurement applicable |
| 3 | André Filipe Paulino da Silva Bento (2021) [3] | High-intensity interval training in high-school physical education classes:  Study protocol for a randomized controlled trial | No result measurement applicable |
| 4 | ORL Farley (2016) [4] | Five Weeks of Sprint and High Intensity Interval Training Improves Paddling Performance in Adolescent Surfers | There is no blank control |
| 5 | Vivian Bertoni Xavier (2020) [5] | Combined aerobic and resistance training improves respiratory and exercise  outcomes more than aerobic training in adolescents with idiopathic scoliosis: a randomized trial | There is no blank control |
| 6 | Leskinen (2011) [6] | [This is an electronic reprint of the original article. This reprint may differ from the original in pagination and typographic detail.](http://research.aalto.fi/files/26124425/paper7.pdf) | No result measurement applicable |
| 7 | Víctor Hugo Arboleda-Serna (2019) [7] | Effects of high-intensity interval training compared to moderate-intensity continuous training on maximal oxygen consumption and blood pressure  in healthy men: A randomized controlled trial | Age out of range |
| 8 | TA Bakken (2013) [8] | Effects of block periodization training versus traditional periodization training in trained cross-country skiers | No result measurement applicable |

6.Leskinen, P Hyvönen, Tuominen E , et al. This is an electronic reprint of the original article. This reprint may differ from the original in pagination and typographic detail.

7.Víctor, Hugo, Arboleda-Serna, et al. Effects of high-intensity interval training compared to moderate-intensity continuous training on maximal oxygen consumption and blood pressure in healthy men: A randomized controlled trial[J]. Biomedica : revista del Instituto Nacional de Salud, 2019, 39(3):524-536.

8.Bakken T A . Effects of block periodization training versus traditional periodization training in trained cross country skiers[J]. Department of Sport & Health Sciences, 2013.

**File 3** Detailed table of basic characteristics of included studies.

| **Group**  **Characteristics** |  | **HIIT Group** | **Control Group** | **Total** |
| --- | --- | --- | --- | --- |
| **Gender**  **(Unit: person)** | Boys | 741(42.37%) | 424(34.03%) | 1165(38.90%) |
|  | Girls | 686(39.22%) | 470(37.72%) | 1156(38.60%) |
|  | Only Girls | 163(9.32%) | 163(13.08%) | 326(10.88%) |
|  | Only Boys | 107(6.12%) | 107(8.59%) | 214(7.15%) |
|  | Gender-neutral | 322(18.41%) | 374(30.02%) | 696(23.24%) |
|  | Total | 1749 | 1246 | 2995 |
| **Weight (Overweight/obesity)**  **(Unit: person)** | Boys | 84(37.50%） | 70(32.71%) | 154(35.16%) |
|  | Girls | 92(41.07%) | 84(39.25%) | 176(40.18%) |
|  | Only Girls | 81(31.16%) | 75(35.05%) | 156(35.62%) |
|  | Only Boys | 56(25%) | 107(8.59%) | 163(37.21%) |
|  | Gender-neutral | 109(27.81%) | 60(28.04%) | 169(38.58%) |
|  | Total | 224 | 214 | 438 |
| **Follow-up Time (Unit: term)**  **Average Weeks：10.99 Weeks** | 1.57Weeks(10Days)/40Weeks | 1(2.13%) /1(2.13%) | | |
|  | 8 Weeks | 14（29.79%) | | |
|  | 12 Weeks | 10（21.82%） | | |
| **Exercise Frequency**  **(Unit: term)** | Twice/ Weeks | 18(40.00%) | | |
|  | 3 times/ Weeks | 21(46.67%) | | |
|  | 4 times / Weeks | 2(4.44%) | | |
|  | 5 times / Weeks | 1(2.22%) | | |
|  | Increase exercise frequency and not explicitly described | 5(11.11%) | | |
|  | Total | 45 | | |
| **Exercise Mode (Unit: term)** | Power bike | 3(6.38%) | | |
|  | Running (indoor/outdoor) | 19(40.43%) | | |
|  | Else | 21(44.68%) | | |
|  | Not explicitly described | 5(8.51%0 | | |
| **Age（5～19）** | Children (5~15) | 2328(77.73%) | | |
|  | Adolescents(＞15) | 667(22.27%) | | |
| **Participants (Unit: person)** | Overweight and Obese | 438(14.62%) | | |
|  | Athletes | 62(2.07%) | | |
|  | Sick children and adolescents | 261(8.71%) | | |
| **Medical Supervision**  **(Unit: term)** | With medical supervision | 26 | | |
|  | Without medical supervision | 0 | | |
|  | Not explicitly described | 21 | | |

**File 4** Measurement method(s) and distribution of outcome indicators.

| Sorts | Outcomes | Measurement method(s) |
| --- | --- | --- |
| Physical morphology | BMI  (37) | digital scale measurements [38, 65-72, 74, 75, 77-82, 84-90, 92, 93, 95-97, 100, 101, 103-106, 108] |
|  |  | not explicitly described [65] |
|  | BF%  (20) | multi-frequency bioelectrical impedance technology body composition analyzer [66, 70, 72, 77, 82, 95-99, 105] |
|  |  | skinfold thickness calculation method [63, 64, 67, 73, 74, 103] |
|  |  | Dual Energy X-ray Absorptiometry (DEXA)[38, 87] |
|  |  | not explicitly described [68] |
|  | WC  (16) | tape measure [66, 78, 87, 89, 90, 97, 98, 103-105, 108] |
|  |  | not explicitly described [38, 82, 95, 99, 100] |
| CRF | VO_2max_  (19) | power bikes, treadmills and other external equipment  [67, 72, 76, 82, 87, 90, 91, 95] |
|  |  | portable cardiopulmonary function tester [38, 63, 65, 97, 99] |
|  |  | 20m turn-back running method [79, 80, 85, 89, 94, 104] |
|  | SBP/DBP  (14/14) | automatic blood pressure tester [38, 83, 89, 104, 105] |
|  |  | manual blood pressure tester [66, 70, 95, 98] |
|  |  | not explicitly described [68, 72, 75, 100] |
|  | HR_max_  (11) | heart rate monitor [63, 65, 67, 70, 82, 86, 91, 92, 95, 98, 102] |
|  |  | not explicitly described [84] |
| Cardiometabolic indexes | TC/TG/HDL-C  (8/8/7) | enzymatic methods [95, 97] |
|  |  | standard colorimetric assays [90] |
|  |  | spectrophotometry [89] |
|  |  | portable measuring instrument [100] |
|  |  | not explicitly described [68] |
|  | LDL-C  (7) | Friedewald formula [63, 73, 80, 89, 92] |
|  |  | portable measuring instrument [95] |
|  |  | not explicitly described [97] |

**Supplemental figures:**

**Fig. 1** Sensitivity analysis of BMI.

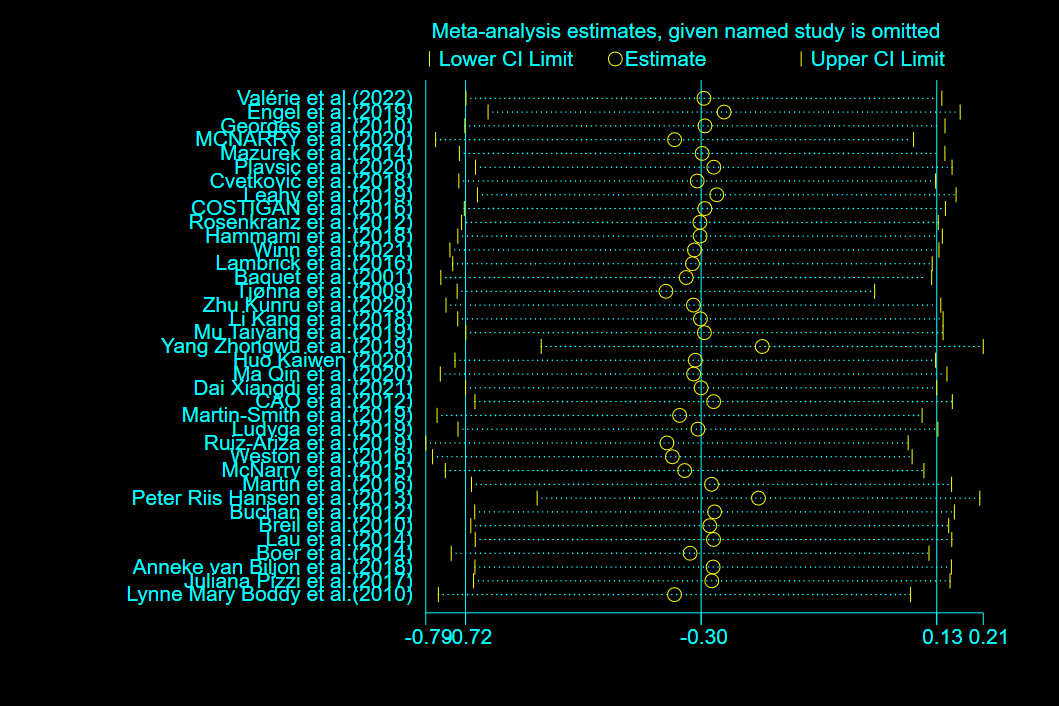

**Fig. 2** Sensitivity analysis of BF%

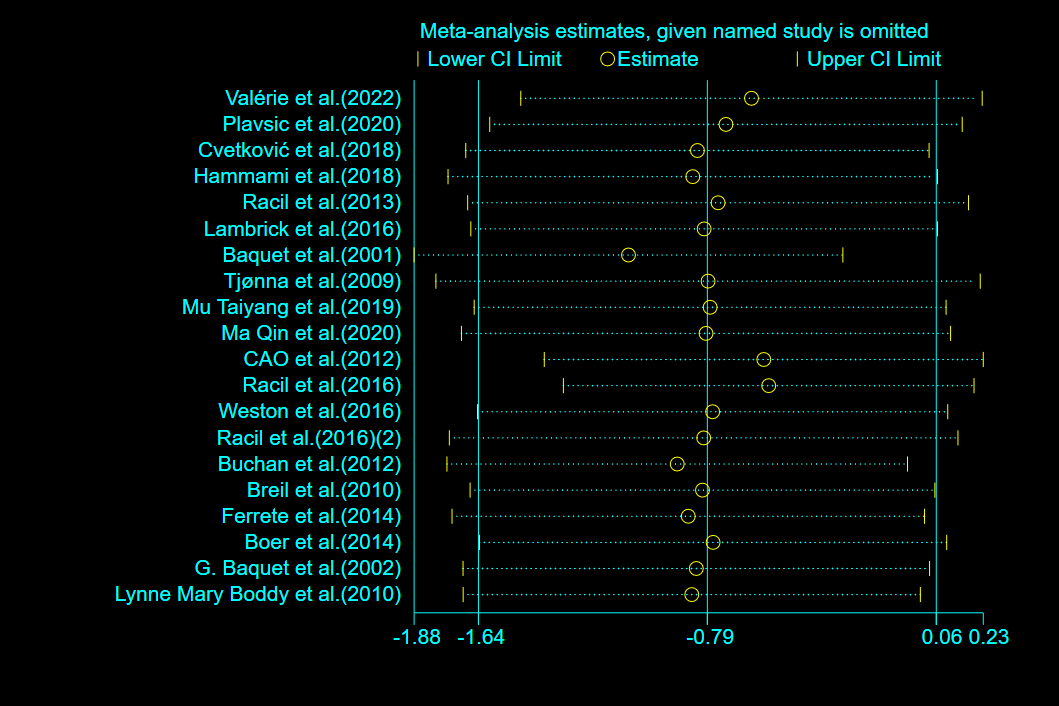

**Fig. 3** Sensitivity analysis of WC

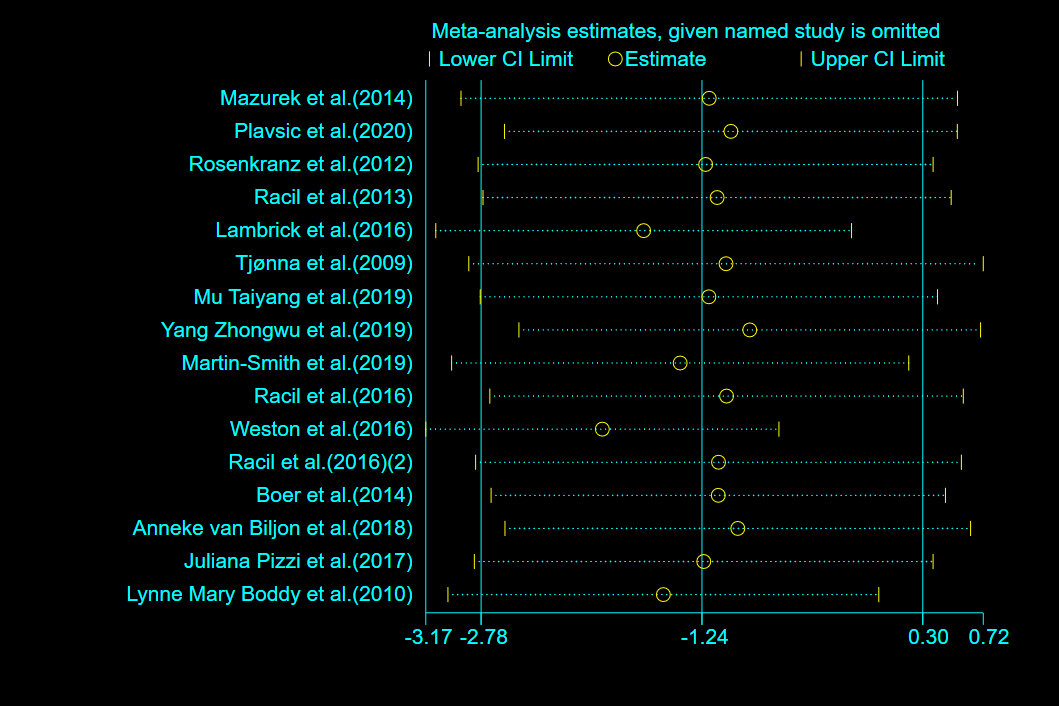

**Fig.4** Sensitivity analysis of TC

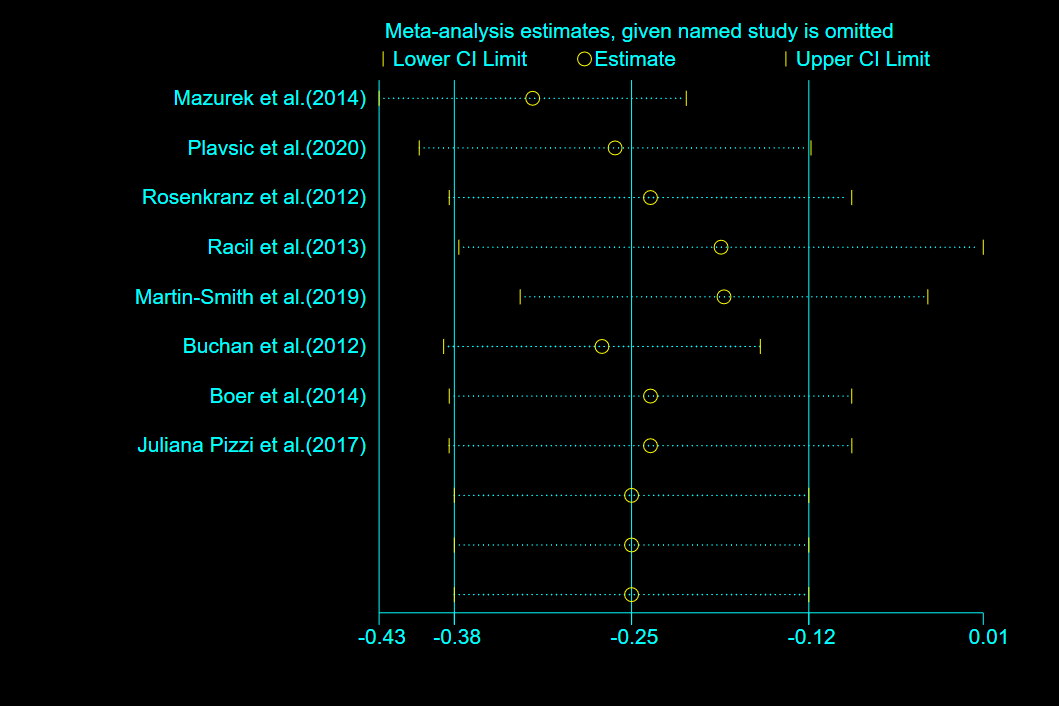

**Fig. 5** Sensitivity analysis of TG

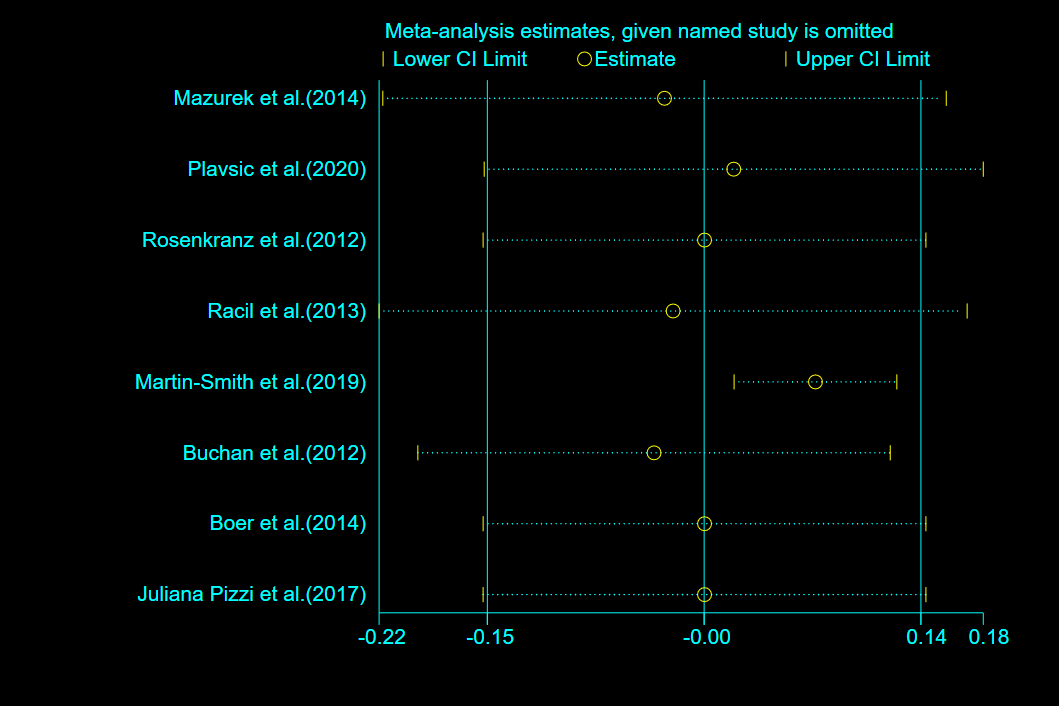

**Fig. 6** Sensitivity analysis of HDL-C

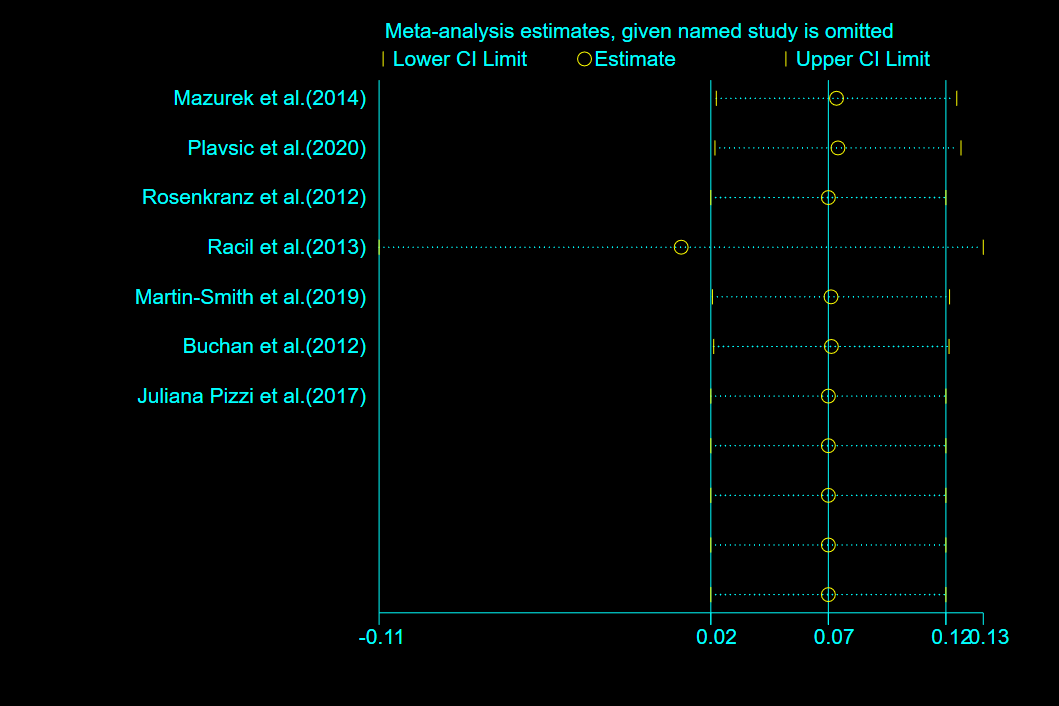

**Fig. 7** Sensitivity analysis of LDL-C

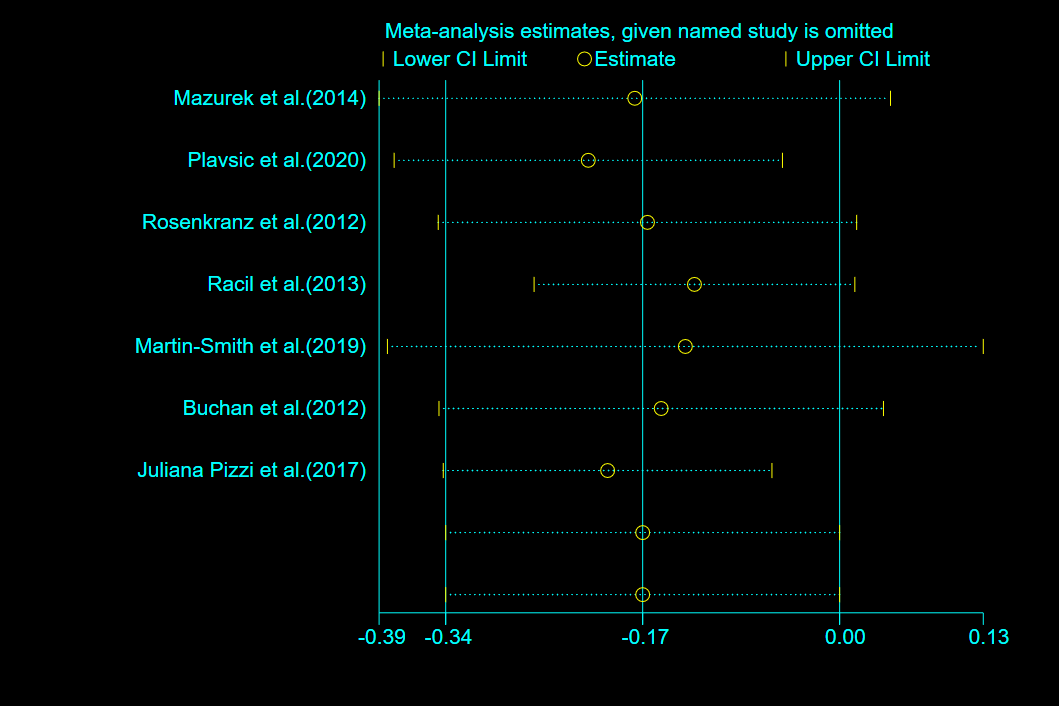

**Fig. 8** Sensitivity analysis of VO_2max_

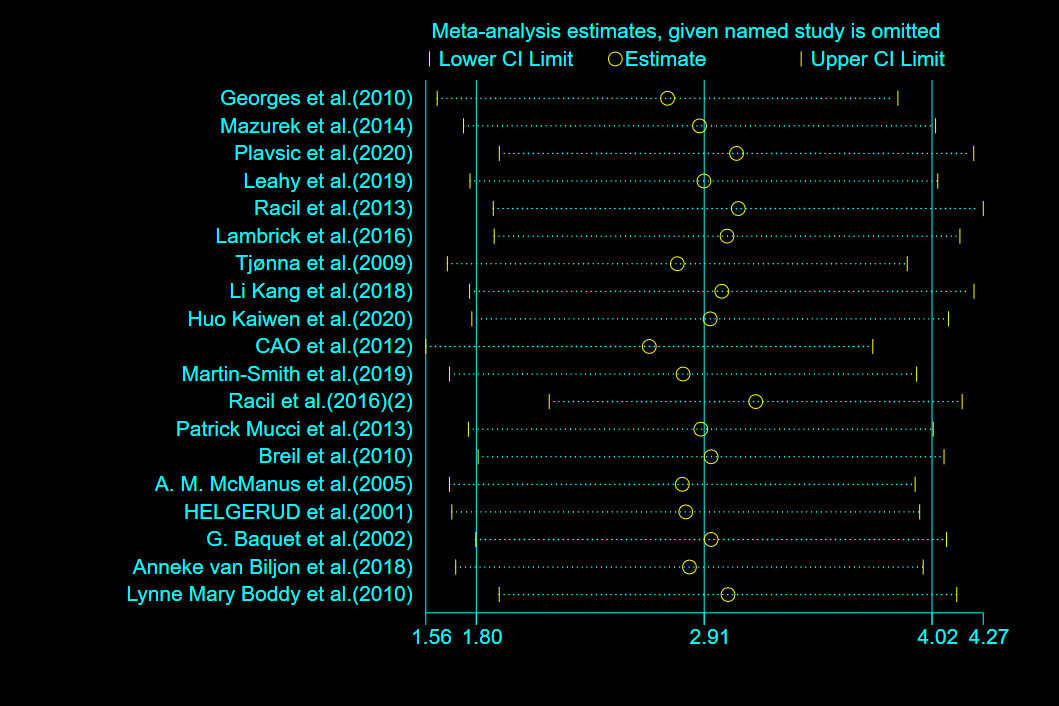

**Fig. 9** Sensitivity analysis of SBP

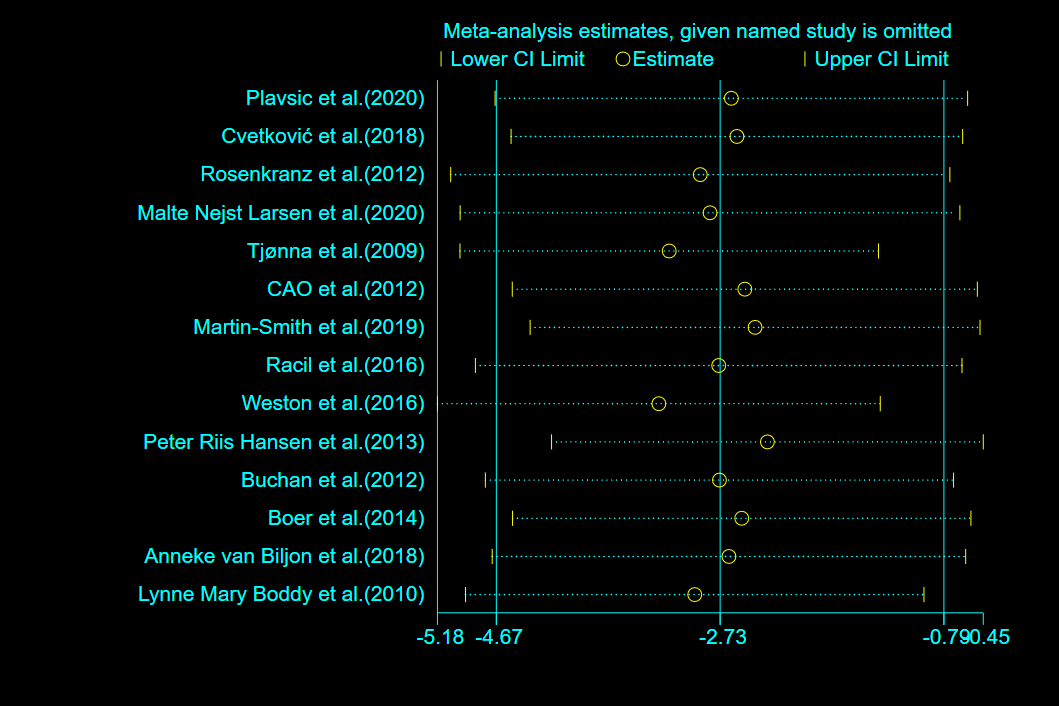

**Fig. 10** Sensitivity analysis of DBP

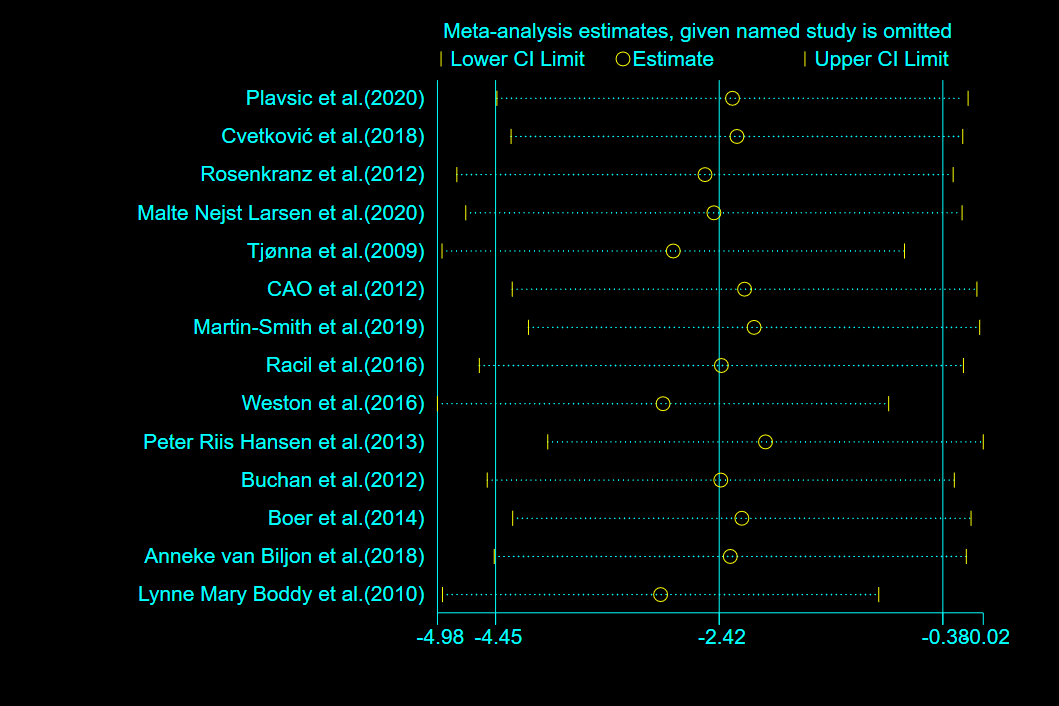

**Fig. 11** Sensitivity analysis of HR_max_

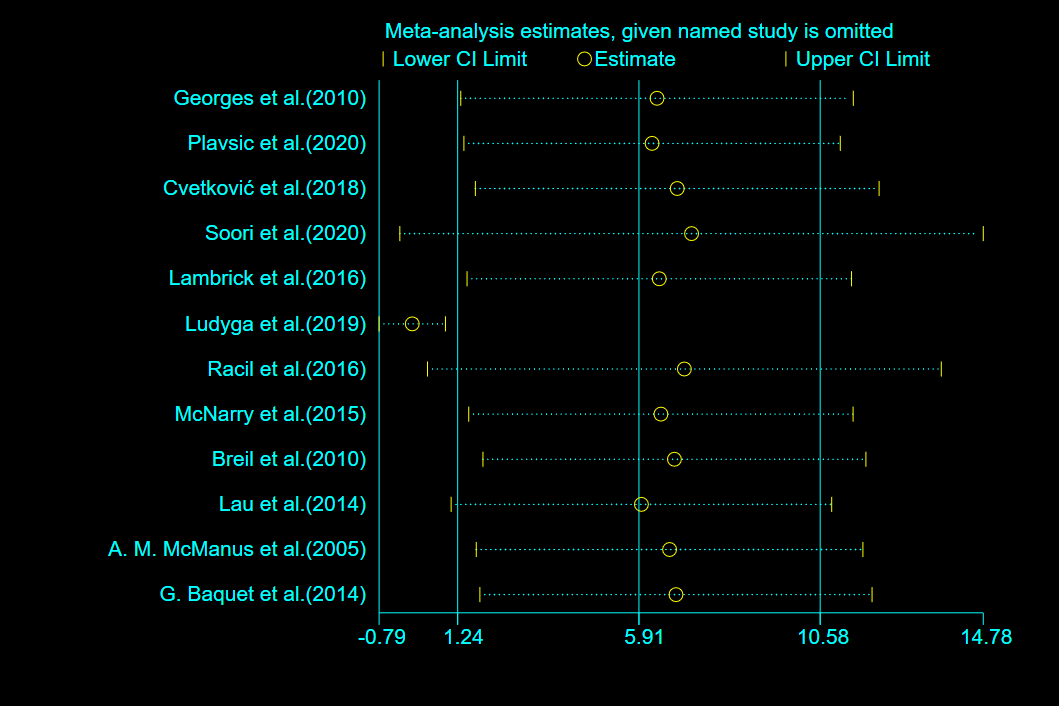
